# Adverse Childhood Experiences and Weight Loss in Overweight and Obese Children in a 9-Year Study: A Prospective Cohort Study with Structural Equation Modeling

**DOI:** 10.1101/2025.07.18.25331809

**Authors:** Seerat Waraich, Hannah Steiman De Visser, Brenden Dufault, Jonathan McGavock

## Abstract

**Background:** Exposure to adverse childhood experiences (ACEs) is associated with a 30 to 50% increased risk of obesity in adolescence. The role of ACEs as a determinant of weight loss among overweight and obese children remains unclear.

**Methods:** Among 8568 nine-year-old children randomly sampled in 2007/2008 for the Growing up in Ireland cohort, 2210 were overweight or obese at 9 years and provided complete follow-up data at age 13 and 18 years. Structural equation and natural effects mediation models tested for a direct causal relationship between ACEs before 9 years and remission risk at 18 years, and indirect effects mediated via daily activity, diet quality, self-image and behavioural difficulties and BMI at 9 years.

**Results:** Among the 1676 adolescents that were overweight or obese at age 9, 46% achieved healthy weight status by age 18; 56% (n=618) of overweight children and 27% (n=153) of obese children), 13% experienced an ACE, and 41% were female. Exposure to an ACE was associated with a higher BMI Z at ages 9 (0.47 vs 0.36, p < 0.05) and 13 years (0.39 vs 0.29, p < 0.05) and reduced the odds of achieving healthy weight status at age 18 by 27% (OR: 0.73, 95% CI:0.54- 0.99). Overweight and obese children exposed to an ACE had lower household income, higher behavioural difficulties, and lower self-concept at ages 9, 13 and 18. Children exposed to an ACE were also 2 to 3-fold more likely to have started smoking before 12 years old and 50% more likely to smoke more than 10 cigarettes a week regularly vape. Behavioural difficulties, self-concept and baseline weight status, but not smoking or dietary habits, mediated the association between ACE exposure and achieving healthy weight at age 18.

**Conclusion:** Among overweight and obese children, exposure to ACEs indirectly reduces the likelihood of achieving healthy weight status at age 18, mediated by its effects on weight in childhood, and behavioural difficulties and self-concept in mid-adolescence. These findings highlight the complex factors that influence weight loss among overweight and obese children exposed to adverse experiences early in life.

## Introduction

Nearly one third of adolescents in Western countries live with overweight or obesity.^1,2^ Clinical practice guidelines recommend intensive behavioural therapy to support weight management in children and adolescents (referred to herein as youth) living with obesity.^3–6^ Despite decades of randomized controlled trials testing various weight management strategies, weight loss among youth living with obesity remains challenging.^6,7^ Specifically, intensive behavioural interventions elicit modest weight loss (0.5-2kg),^7,8^ with significant inter-individual variation in both the response to interventions^9^ and weight trajectories throughout adolescence.^10–12^ Established sources of resistance to and variation in weight loss among overweight and obese youth include baseline fitness,^9^ household income,^13^ and age.^14^ While psychosocial factors^15^ and adverse childhood experiences (ACEs)^16–18^ are established determinants of pediatric obesity, their role as determinants of weight loss in overweight and obese youth remains unclear.

To determine the influence of exposure to ACEs and associated psychosocial consequences on weight loss among overweight and obese youth, we applied causal modeling to a large population-based cohort followed prospectively for 9 years. The primary study hypothesis guiding this study was that exposure to ACEs in childhood is associated with a reduced likelihood of weight loss by age 18, compared to children unexposed, and that this association would be mediated by behavioural and emotional factors in mid-adolescence (Figure 1).

**Figure 1.**
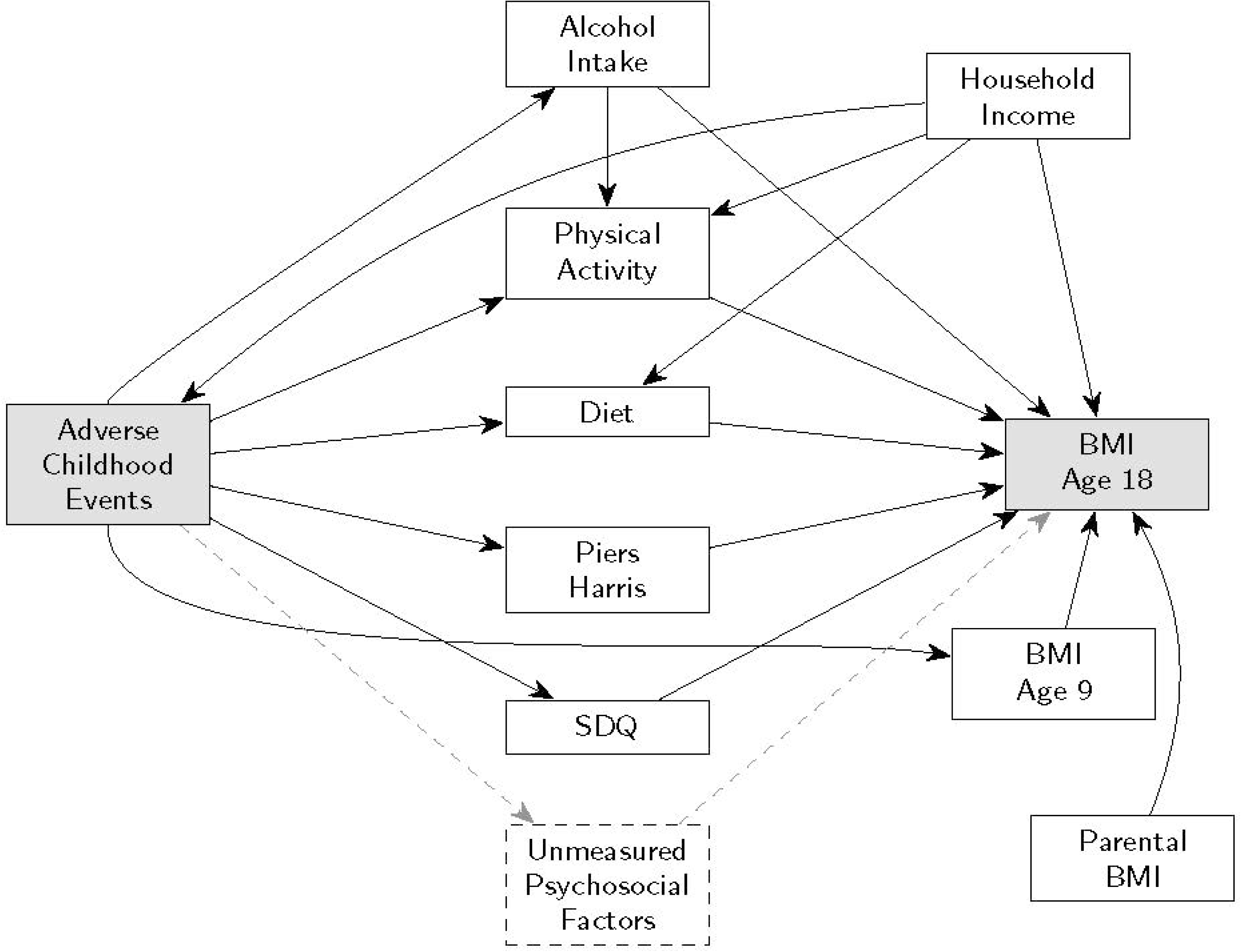
Directed acyclic graph (DAG) describing possible pathways through which exposure to an adverse childhood experience among overweight and obese children could influence achieving healthy weight status by age 18 years.

## DESIGN AND METHODS

### Study Population, Design and Sampling

To test the study hypothesis, we used data from the Growing up in Ireland Child Cohort study that included 8568 9-year-old children and their families, followed until age 18.^20,21^ Data collection visits occurred when the child was 9 (2007-2008), 13 (2012-2013) and 18 years of age (2015-2016), with 6039 child-caregiver pairs having data in all three time points. For the proposed study, we restricted analyses to the 2210 children considered overweight or obese according to their sex-specific BMI Z score at age 9. There were no exclusion criteria for cohort entry. Parents consented and children assented to the study during home visits. All stages of the Growing Up in Ireland project were approved by the Health Research Board’s standing Research Ethics Committee based in Dublin, Ireland. Data for the current study were accessed via the Irish Social Science Data Archive (https://www.ucd.ie/issda/).

### Primary Exposure

The primary exposure of interest were adverse experiences in childhood before age 9, treated as a binary variable. Exposure to an adverse experience was defined as responding “yes” to any of 14 adverse experiences by the primary caregiver during the household visit when the child was 9 years old.^16^ An additional exposure was restricted to the four adverse experiences included in the original ACEs study: divorce, parent in prison, parent with drug/alcohol abuse, and parent with a mental health disorder.^22,23^ Children exposed to at least one of these four adverse experiences are classified as being exposed to an ACE. The other ten experiences were adapted from the National Longitudinal Survey of Children and Youth.^24,20^ Children whose parents reported “none” were treated as unexposed to any adversity.

### Primary outcome

The outcome measure was weight status at age 18, treated as a binary variable and classified as either normalization of body weight: < 25 kg/m^2^; or remaining overweight or obese (≥ 25.0kg/m^2^). Each young person’s height and weight were measured objectively by a trained interviewer in the adolescent’s home at 9, 13 and 18 years of age. BMI classifications of non-overweight, overweight, or obese, at ages 9 and 13, were determined using age and sex-specific Z scores developed from international data by the World Obesity Federation.^25,26^

### Mediating Variables of Interest

Behavioural difficulties were measured using the strengths and difficulties questionnaire (SDQ)^27,28^ and was completed by the primary caregiver when the child was aged 9, 13 and 18. The SDQ has 5 subscales, including 4 areas of difficulties: emotional, conduct, hyperactivity, peer problems, as well as a prosocial subscale.^27,28^ Subscales were used to calculate a total SDQ score, in which a higher total score indicated more behavioural difficulties. Self-concept was measured using the Piers-Harris questionnaire,^29^ completed by the child at 9 and 13. The Piers- Harris questionnaire is a 60-item self-reported questionnaire that assesses self-concept through six different subscales: behavioural adjustment, intellectual and school status, physical appearance and attributes, freedom from anxiety, popularity, and happiness and satisfaction.^29^ A higher score on the Piers-Harris indicated a higher sense of self-concept. For the main structural equation model (SEM), behavioural difficulties (SDQ total score) and self-concept (Piers Harris Total Score) at age 13 were treated as mediators of the association between ACE exposure and healthy weight status at age 18 years.

### Covariates and Confounding

Potential confounders and covariates collected when the child was 18 years old were used in the SEM and were chosen based on previously documented associations with weight status in childhood or adolescence.^30–32^ Potential confounders of the relationship between adverse childhood events and normalization of body weight included household income and household social class. Household income was self-reported and normalized to income per household member. Other co-variates included primary caregiver BMI. Primary caregiver BMI was measured objectively during the home visit that occurred when the adolescent was 18 years of age.

Self-reported dietary habits and physical activity were measured at ages 9, 13 and 18. A diet quality score was calculated based on a previous study.^33^ Questions about how often the adolescent had consumed a type of food in the past 24 hours were used. For foods classified as healthy, a score of 2 was given if consumed more than once, 1 if consumed once, and 0 if not consumed at all. Foods classified as unhealthy were given a score of -2 if consumed more than once, -1 if consumed once, and 0 if not consumed at all. The scores of each of the different food categories were summed to calculate the total diet quality score. To estimate daily physical activity, adolescents reported the number of times in the last 14 days that they accumulated at least 20 minutes of exercise; used as a proxy for habitual physical activity.

### Statistical Methods

A series of T-tests and Chi-Square tests were used to test for differences in demographic variables, mediators and outcomes for overweight and obese youth exposed at ages 9, 13 and 18 years. A logistic regression model was used to compare the odds of normalizing body weight at age 18 yearsfor overweight and obese youth exposed to an ACE, compared to those unexposed, adjusting for household income. This model gives the adjusted total effect of ACE on BMI remission. Our mediation hypotheses decompose this total effect into candidate pathways and were tested with a structural equation model (SEM) and natural effects model. The variables included in the full SEM were determined from the a-priori causal model between ACEs and weight loss in adolescence (Figure 1). Two of the mediating variables, behavioural difficulties and self-concept, were treated as latent variables. Variables used to estimate these latent variables are provided in eTable 1. This incorporates the measurement properties of the two scales directly into the overall estimation of the SEM. We assumed income was a common cause of ACEs and BMI at all three time points and, therefore, a confounder, and that BMI at age 9 years and parental BMI would be associated with a child’s likelihood of normalizing body weight by age 18 years. Mediation effects were calculated for the indirect and total association between exposure to any adverse experience, self-concept, behavioural difficulties, and BMI at age 18 years. The magnitude of the mediation is calculated as the product of the estimated standardized paths between an exposure (any adversity/ACE), a mediator (behavioural difficulties, self-concept, physical activity, dietary patterns, smoking/vaping) and the outcome of interest (normalization of body weight). The total effect of any adversity or ACE on normalization of body weight is calculated as the sum of the direct effect and two mediated effects. These estimates yield valid causal estimates in the absence of unmeasured confounding, non-linearity, and interaction. Diagonally-weighted least squares (DWLS) were used for all SEMs because the exposure and outcome, ACES and normalization of body weight, were ordinal. When using DWLS as the estimation method, the structural (path) estimates are based on the probit regression model. This assumes a normal latent distribution underlying the observed categorical variables.

An important caveat within our proposed causal model was the potential causal relationship between two mediators of interest: alcohol intake and daily physical activity. Smoking behaviours were not included due to large missingness. As posed in our causal diagram, alcohol intake may be a cause of physical activity; specifically, increased alcohol use may lower daily physical activity. However, alcohol use plays the dual role of confounder (with respect to physical activity and achieving healthy weight status) and mediator (with respect to ACE exposure and achieving healthy weight status). As the confounder is itself caused by exposure, the indirect effects via physical activity are not generally identifiable, meaning their values cannot be recovered from observable data even with confounder adjustment.^34^ These values are recoverable under certain parametric assumptions like linearity^35^ or fully specified parametric g-estimation.^36^ Instead of taking these approaches, we chose to be agnostic about the exact causal relationship between these mediators, which may include unmeasured common causes (for example, a latent “health attitudes” causing both alcohol and exercise), and chose a natural effects joint mediation approach.^37^ With joint mediation, we treat the indirect mediated effect as being through either alcohol use or physical activity or both, without specifying which particular path. Estimation proceeded using a form of marginal structural models with imputation. Another advantage is that the direct and indirect effects can be expressed on the scale of the outcome variable; odds ratios in our case of BMI remission. The R package lavaan^38^ was used for SEM analyses. The R package medflex was used for the joint mediation analysis.^37^ We used R version 4.3.1 (www.R-project.org) for all data management and statistical modeling. The R code used for all analyses is provided in the appendix.

## RESULTS

### Participant characteristics

A flow chart describing how the final sample size was achieved is provided in eFigure 1. Of the original cohort of 8568 9-year-old children, 6216 children returned for follow-up at age 18. Of these 6216 children, 2022 were overweight or obese at age 9 and 1676 had data for all three years and were included in the analysis. At age 9, children exposed to an ACE displayed higher BMI Z scores, lower self-concept and greater behavioural difficulties, without any differences in self-reported physical activity or diet quality (Table 1). Primary caregiver depression scores were nearly 2-fold higher among ACE-exposed children than those unexposed. At all three time points, ACE-exposed children lived in households with lower income.

**Table 1.** Demographic, mediators, and outcomes between overweight and obese youth exposed and unexposed to an adverse childhood experience (ACE) at age 9 years.

| **Variables** | **No ACE**<br>**(n=1461)** | **ACE**<br>**(n=215)** | **P-value** |
| --- | --- | --- | --- |
| BMI Z score | 1.81 (0.56) | 1.92 (0.62) | 0.0279 |
| Boys (n / %) | 755 (51.7%) | 91 (42.3%) | 0.0106 |
| Household Income (1000<br>€/person) | 21.4 (14.1) | 18.3 (11.9) | <0.001 |
| Hard Exercise last 14 days |  |  |  |
| 4 to 5 days | 316 (21.6%) | 50 (23.3%) |  |
| 5+ days | 728 (49.8%) | 106 (49.3%) | 0.60 |
| Diet Score | 2.00 (3.70) | 1.71 (3.93) | 0.346 |
| Self-Concept Score | 46.8 (8.31) | 45.0 (9.86) | 0.06 |
| Behavioural Diffs Score | 7.48 (4.86) | 9.61 (5.95) | <0.001 |
| Primary Caregiver Variables |  |  |  |
| BMI (kg/m<sup>2</sup>) | 27.3 (4.95) | 26.9 (4.58) | 0.443 |
| Depression Score | 1.77 (2.85) | 3.41 (4.65) | <0.001 |

Data are provided as mean (SD) and count (%).

### Behavioural consequences of ACE exposure for overweight and obese children

Compared to unexposed youth, those with ACE exposure were 25% more likely to have smoked before ever, 3-fold more likely to have started smoking before age 12, smoked significantly more cigarettes weekly, and were 33% more likely to have tried vaping (Table 2). ACE exposure before 9 years was also associated with lower subscales of self-concept and higher subscales of behavioural difficulties and by extension lower self-concept (β = -0.097; 95% CI: -0.12 to - 0.073) and behavioural difficulty (β = +0.17; 95% CI: +0.14 to +0.21) total scores at age 13 years. At ages 13 and 18, parents of youth exposed to an ACE displayed higher levels of depression and stress scores, compared to parents of unexposed youth (Tables 3 and 4). By age 18, overweight and obese youth exposed to an ACE displayed lower levels of emotional stability, lower problem-solving and higher avoidance scores for coping strategies and lower self-control. No differences were observed in other measures of personality subscales (Table 4).

**Table 2.** Select lifestyle behaviours and secondary exposure to ACEs of overweight and obese children at age 13 years, stratified based on exposure to an adverse experience before 9 years old.

| **Variable** | **No ACE**<br>**(n=1461)** | **ACE**<br>**(n=215)** | **P-value** |
| --- | --- | --- | --- |
| Hard Exercise Last 14 days |  |  |  |
| 4-5 times | 283 (19.4%) | 24 (11.2%) | 0.007 |
| >5 times | 287 (19.6%) | 34 (15.8%) |  |
| Tried smoking ever | 688 (47.7%) | 128 (59.8%) | 0.001 |
| Tried smoking before 13 years | 48 (7.05%) | 18 (14.1%) | 0.013 |
| >5 Cigarettes/Week | 46 (18.0%) | 30 (40.0%) |  |
| Tried Vaping ever | 463 (32.1%) | 91 (42.5%) | 0.003 |
| Ever tried Alcohol | 1303 (90.4%) | 195 (91.1%) | 0.804 |
| **Secondary exposure to adverse events by 13 years** |  |  |  |
| Divorce | 91 (6.23%) | 56 (26.4%) | <0.001 |
| Violence at home | 117 (8.01%) | 39 (18.4%) | <0.001 |
| Lost best friend | 100 (6.84%) | 24 (11.3%) | 0.03 |

Data are provided as count (%).

**Table 3.** Behavioural difficulties, self-concept and caregiver depression at 13 years of age among overweight and obese children stratified by ACE exposure.

| **Variables** | **No ACE**<br>**(n=1461)** | **ACE**<br>**(n=215)** | **P-value** |
| --- | --- | --- | --- |
| **Self-Concept Sub scales** |  |  |  |
| Behaviour | 12.7 (1.97) | 12.2 (2.28) | <0.001 |
| Intellect | 12.3 (2.96) | 11.3 (3.41) | <0.001 |
| Physical | 7.79 (2.40) | 7.20 (2.75) | 0.008 |
| Free from anxiety | 10.5 (3.10) | 9.69 (3.55) | 0.002 |
| Popularity | 9.63 (2.29) | 9.24 (2.61) | 0.06 |
| Happiness | 8.45 (1.73) | 7.95 (2.15) | 0.005 |
| Total Score | 47.7 (8.33) | 45.0 (10.1) | <0.001 |
| **Behavioural Difficulties** |  |  |  |
| Emotional regulation | 1.75 (1.94) | 2.52 (2.14) | <0.001 |
| Conduct | 1.00 (1.30) | 1.45 (1.77) | <0.001 |
| Hyperactivity | 2.40 (2.23) | 3.11 (2.63) | <0.001 |
| Peer interactions | 1.12 (1.48) | 1.68 (1.86) | <0.001 |
| Prosocial Behaviour | 8.96 (1.45) | 8.68 (1.62) | 0.0147 |
| Total Score | 6.28 (4.89) | 8.76 (6.14) | <0.001 |
| **Parental Depression** |  |  |  |
| PCG Depression | 2.13 (3.01) | 4.31 (5.38) | <0.001 |
| SCG Depression | 1.49 (2.52) | 2.22 (3.45) | 0.392 |
| Missing | 268 (18.3%) | 141 (65.6%) |  |

Data are provided as mean (SD) and count (%). PCG = primary care giver; SCG = secondary care giver; ACE = adverse childhood experience.

**Table 4.** Select behavioural outcomes at age 18 years among overweight and obese children stratified by ACE exposure..

| **Variables** | **No ACE**<br>**(n=1461)** | **ACE**<br>**(n=215)** | **P-value** |
| --- | --- | --- | --- |
| **Ten Item personality scale** |  |  |  |
| Extravert | 4.81 (1.36) | 4.69 (1.35) | 0.305 |
| Agreeable | 4.70 (1.01) | 4.70 (1.14) | 0.777 |
| Conscientious | 5.20 (1.13) | 5.12 (1.25) | 0.391 |
| Openness | 5.49 (1.02) | 5.63 (0.96) | 0.08 |
| Emotional stability | 4.77 (1.34) | 4.46 (1.48) | 0.003 |
| **Level of self-control** | 3.40 (0.687) | 3.21 (0.728) | <0.001 |
| **Coping strategy indicator** |  |  |  |
| Problem solving subscale | 16.6 (5.10) | 15.9 (4.86) | 0.0446 |
| Avoidance subscale | 13.5 (5.46) | 15.4 (6.05) | <0.001 |
| Rosenberg self-esteem score | 12.0 (3.50) | 10.9 (3.98) | <0.001 |
| Internal locus of control | 23.8 (3.08) | 24.0 (3.47) | 0.252 |
| Prosocial Behaviour | 8.96 (1.45) | 8.68 (1.62) | 0.0147 |
| **Parental Health** |  |  |  |
| Total depression score | 2.43 (3.35) | 4.23 (5.07) | <0.001 |
| Parental stress score | 10.1 (3.71) | 11.9 (4.34) | <0.001 |
| Equivalized Household annual income | 16.3 (9.02) | 13.7 (6.19) | <0.001 |

Data are provided as mean (SD)

### ACE Exposure and the likelihood of achieving healthy weight status at age 18 years

Overweight and obese children exposed to an ACE were 27% less likely (OR: 0.73; 95% CI: 0.54-0.99) to achieve healthy weight status at age 18 compared to those unexposed. In addition to exposure to an ACE before age 9, regression models revealed that daily exercise, self-concept, behavioural difficulties and baseline BMI were also associated with the likelihood of achieving healthy weight status by age 18 (eTable 2). Some of these associations remained significant when analyses were repeated using a structural equation model (eTable 3). Within the final structural equation model, after adjusting for household income, standard path co-efficients revealed that BMI Z score at age 9, behavioural difficulties (β = -0.022; 95% CI: -0.05 to +0.001) and self-concept (β = +0.01; 95% CI: +0.002 to -0.018) at age 13 years mediated the association between exposure to an ACE and the reduced likelihood of achieving healthy weight status at 18 years (eTable 4, Figure 2). Dietary score and daily physical activity were not significant mediators of the association between ACE exposure and achieving healthy weight status at 18 years (Figure 2). Natural effects joint mediation revealed that there was no joint mediation between alcohol intake and physical activity for the association between ACE exposure and achieving healthy weight status at 18 years (natural indirect effect OR 0.98; 95% CI: 0.93 – 1.03) (eTable 5).

**Figure 2.**
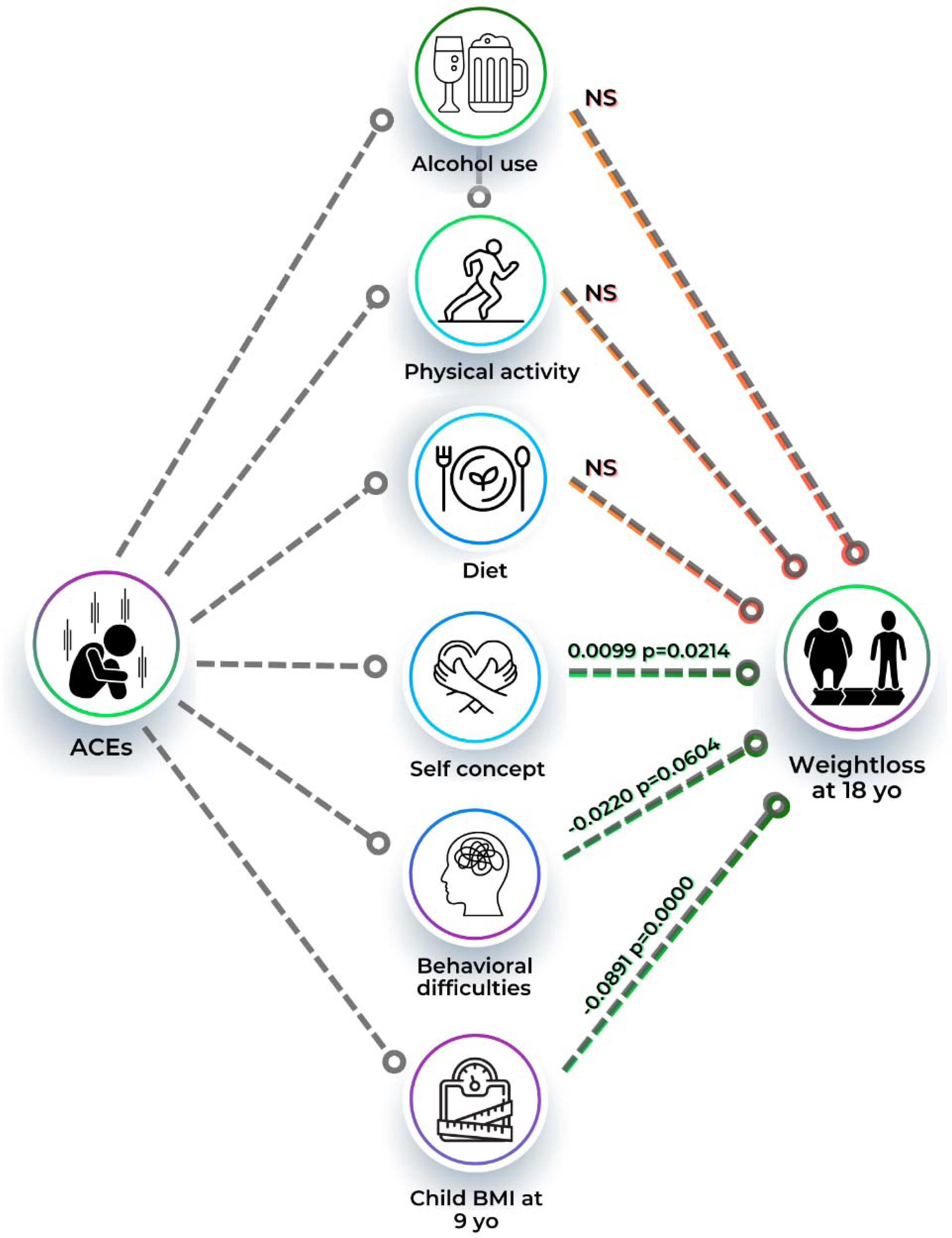
Structural equation model describing the results from the structural equation model testing for a causal effect of exposure to an ACE and achieving a healthy weight status by 18 years of age.

## DISCUSSION

The main finding from this novel causal approach to studying the determinants of weight loss in overweight and obese children were that achieving healthy weight status is reduced nearly 27% for those exposed to an ACE before age 9 years. Second, we found that overweight and obese children exposed to an ACE before age 9 years are more likely to engage in several unhealthy behaviours like smoking and vaping by age 13, and their parents display higher depression and stress than those of unexposed children. Additionally, ACE exposure before age 9 years was associated with poorer coping strategies, self-control and emotional stability at age 18 years. Lastly, structural equation models revealed that weight status at age 9 years, lower self-concept, and higher behavioural difficulties at age 13 years indirectly mediate the association between exposure to both ACEs and achieving healthy weight status by age 18 years among overweight and obese children.

Adolescent obesity is a complex chronic disease, and weight loss among adolescents is challenging.^2,15^ Systematic reviews of intervention studies reveal modest weight loss following intensive behavioural therapy interventions,^6,7^ with significant inter-individual variation.^39,40^ Applying novel causal modelling methods to a large cohort of overweight and obese youth, we expand on results from experimental trials with smaller sample sizes by demonstrating that a complex set of social and behavioural factors are also determinants of weight loss throughout adolescence. We found significantly higher levels of behavioural difficulties, difficulties with emotional regulation and coping strategies, as well as a higher prevalence of unhealthy behaviours, which could influence behaviour change among overweight and obese children exposed to an ACE early in life. Additionally, we found that higher behavioural difficulties and low self-concept but not diet nor physical activity in mid-adolescence mediate the effect of ACE exposure on the likelihood of achieving healthy weight status at age 18. The mediating role of these psychological consequences of ACE exposure is nearly identical to the effects seen for the role of ACEs as determinants of ischemic heart disease in adults.^41^ While the effect sizes presented here are small, they are quite similar to those observed following large public health interventions aimed at addressing childhood obesity,^42,43^ suggesting relevance from a public health standpoint. These data reinforce the complex nature of adolescent weight management and support tailored approaches to behaviour change.

Exposure to ACEs is associated with an array of behavioural and biological factors that influence lifelong health.^44^ Previous studies have documented dose-response effects of ACE exposure on adolescent mental well-being, emotional status and cardiometabolic health outcomes.^19,44^ To the best of our knowledge, this is the first prospective cohort study documenting that exposure to an ACE reduces the likelihood of achieving healthy weight status among overweight and obese children. Similar to other cohorts,^45^ we found that overweight and obese children exposed to ACE are more likely to engage in smoking and vaping, without any differences in daily alcohol consumption, self-reported physical activity or dietary habits.

Additionally, overweight and obese children exposed to an ACE are also more likely to have lower self-image, a reduced ability to self-manage stress, poorer coping strategies and live in a household with lower income. The consequences of ACE exposure were also associated with a higher level of depression and anxiety among primary and secondary caregivers throughout adolescence. Within the larger framework of precision medicine, the data presented here support the possibility that ACEs (1) are a unique determinant of weight loss in adolescents and (2) lead to a clustering of behavioural consequences that require unique weight management strategies. Collectively, these data reinforce the complex nature of pediatric weight management and the need for more person-centred behavioural approaches tailored to their psychosocial needs/exposures.

The study is strengthened by the application of a causal approach to a complex hypothesis with a large, prospective cohort of overweight and obese children followed for nearly a decade. Despite these strengths, there are several limitations that need to be addressed. First, there was significant loss of follow-up with each wave, particularly among families from lower income households. Additionally, not all of the ACEs from the original ACEs study^23^ were included in design, limiting the precision of the exposure to ACEs among children. Additional information, such as the length of exposure, repeated exposures or age at the time of exposure occurred could have influenced the associations observed here and potentially mediate the lasting effects of exposure to ACEs in childhood on weight loss throughout adolescence. Although our model adjusted for various sources of confounding, these findings may also have been influenced by unmeasured confounding.

## CONCLUSIONS

Exposure to any ACEs before age 9 years reduced the odds of achieving healthy weight status by age 18 years, and achieving a healthy weight status by age 18 is mediated more by externalizing behaviours than lifestyle behaviours. ACE exposure before age 9 was also associated with a clustering of behavioural, psychological, structural and parenting factors that could influence the effectiveness of conventional behavioural management interventions aimed at weight loss among overweight and obese children.

## Supporting information

STROBE Checklist

## Data Availability

Data can be made publicly available through an application process through the Canadian Institutes for Health Information.

## Acknowledgements

We are indebted to the families who participated in the Growing Up in Ireland Cohort study and the staff at the Economic and Social Research Institute in Dublin for data collection and management.

