## Supplementary material for "Adverse Childhood Experiences and Weight Loss in Overweight and Obese Children in a 9-Year Study: A Prospective Cohort Study with Structural Equation Modeling": Online Supplement

**Supplemental Information**

eFigure 1. Flow chart for sample size.

eTable 1. Latent variables for the hypothesized mediators self concept and behavioural difficulties.

eTable 2. Results from multivariate linear regression for determinants of achieving healthy weight by 18 years.

eTable 3. Structural Equation Model Regressions Predicting BMI Remission by age 18 and Mediators Using Diagonally Weighted Least Squares.

eTable 4. Standardized Indirect (Mediated) Effects Linking ACE Exposure to Healthy Weight Status at Age 18 Years in Overweight and Obese Children.

eTable 5. Natural Effects Mediation of the Association Between ACE Exposure and Achieving Healthy Weight Status at Age 18 Years via Physical Activity and Alcohol Intake (Adjusted for Household Income).

R code for all analyses.

**eFigure 1. Flow chart for sample size.**

Growing Up in Ireland Child Cohort

(n=8568)

ACE exposure before age 9

(n=215)

No ACE exposure before age 9

(n=1461)

Sample with data for all 3 years and included in SEM

(n=1676)

Excluded (n=346)

Insufficient data across 3 waves.

Excluded (n=2352)

Lost to follow-up by age 18.

Overweight or obese children at age 9

(n=2022)

Children with follow-up data

(n=6216)

**eTable 1. Latent variables for the hypothesized mediators self concept and behavioural difficulties.**

| **Variable** | **Estimate** | **SE** | **Std LV** | **STD All** |
| --- | --- | --- | --- | --- |
| Self Concept |  |  |  |  |
| Physical | 1.94 | 0.05 | 1.96 | 0.79 |
| Free_Anxiety | 2.50 | 0.06 | 2.53 | 0.80 |
| Popularity | 1.60 | 0.04 | 1.62 | 0.69 |
| Happiness | 1.46 | 0.04 | 1.47 | 0.81 |
| Intellectual | 2.41 | 0.06 | 2.44 | 0.79 |
| Behaviour | 0.77 | 0.04 | 0.78 | 0.39 |
| Behavioural Difficulties | |  |  |  |
| Emotional | 1.05 | 0.04 | 1.11 | 0.56 |
| Personal | 0.74 | 0.03 | 0.78 | 0.51 |
| Hyperactivity | 1.20 | 0.05 | 1.27 | 0.55 |
| Conduct | 0.90 | 0.03 | 0.94 | 0.68 |
| Prosocial | -.066 | 0.03 | -0.70 | -0.47 |
| PH behaviour | -0.57 | 0.04 | -0.60 | -0.30 |

**eTable 2. Results from multivariate linear regression for determinants of achieving healthy weight by 18 years.**

| **Variable** | **Estimate** | **SE** | **P value** | **STD lv** |
| --- | --- | --- | --- | --- |
| ACE Exposure | 0.031 | 0.066 | 0.64 | 0.031 |
| Diet Score | -.001 | 0.009 | 0.95 | -0.001 |
| Physical activity | 0.11 | 0.04 | 0.005 | 0.11 |
| Self concept* | -0.07 | 0.027 | 0.014 | -0.067 |
| Behavioural Diff | -0.069 | 0.036 | 0.058 | -0.072 |
| Alcohol Intake | -0.043 | 0.038 | 0.265 | -0.043 |
| Income | 0.004 | 0.002 | 0.112 | 0.004 |
| BMI @ age 9 | -0.187 | 0.016 | <0.001 | -0.186 |

**eTable 3. Structural Equation Model Regressions Predicting BMI Remission by age 18 and Mediators Using Diagonally Weighted Least Squares.**

| **Predictor** | **Std. β** | **Std. Error** | **z-value** | **P-value** |
| --- | --- | --- | --- | --- |
| BMI Z score at 9 | -0.186 | 0.016 | -11.743 | 0.000 |
| Self-concept | -0.067 | 0.027 | -2.468 | 0.014 |
| Behavioural Difficulties | -0.072 | 0.036 | -1.893 | 0.058 |
| Physical activity | 0.108 | 0.039 | 2.789 | 0.005 |
| Diet Score | -0.001 | 0.009 | -0.061 | 0.952 |
| Alcohol intake | -0.043 | 0.038 | -1.114 | 0.265 |
| Equivalized household Income | 0.004 | 0.002 | 1.591 | 0.112 |
| ACE exposure | 0.031 | 0.066 | 0.472 | 0.637 |

β = standardized path coefficient from SEM; DWLS estimator

**eTable 4. Standardized Indirect (Mediated) Effects Linking ACE Exposure to Healthy Weight Status at Age 18 Years in Overweight and Obese Children.**

| **Mediator** | **Std. β** | **Std. Error** | **z-value** | **95% Cl** | **P-value** |
| --- | --- | --- | --- | --- | --- |
| BMI Z score at 9 | -0.089 | 0.015 | -5.996 | -0.118 to -0.060 | < 0.001 |
| Behavioural  Difficulties | -0.022 | 0.012 | -1.882 | -0.045 to 0.001 | 0.060 |
| Self-concept | 0.010 | 0.004 | 2.290 | 0.002 to 0.018 | 0.022 |
| Diet score | 0.000 | 0.002 | 0.060 | -0.004 to 0.004 | 0.952 |

β = standardized path coefficient from SEM; DWLS estimator

**eTable 5. Natural Effects Mediation of the Association Between ACE Exposure and Achieving Healthy Weight Status at Age 18 Years via Physical Activity and Alcohol Intake (Adjusted for Household Income).**

| **Effect Type** | **Mediators Included** | **Odds Ratio** | **95% CI** | **P-value** |
| --- | --- | --- | --- | --- |
| Natural Indirect effect | Physical activity, alcohol intake | 0.98 | 0.93 to 1.03 | 0.36 |
| Natural direct effect | None | 0.82 | 0.60 to 1.12 | 0.21 |
| Total effect | N/A | 0.80 | 0.59 to 1.10 | N/A |

R Code for analyses

GUI: ACE, Mediation, and BMI Remission

Brenden Dufault, MSc

2025-06-06

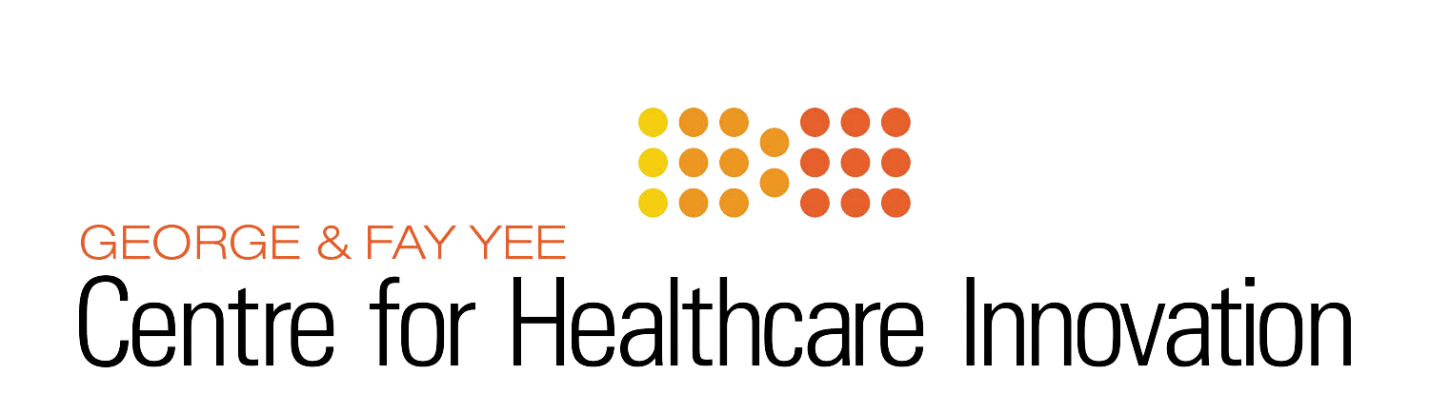

Overview and Important Notes

In this document we investigate whether childhood exposure to ACE at age 9 is causative of BMI remission at age 18 among overweight and obese adolescents, and whether this effect is mediated by a number of psychosocial factors. To this end we employ structural equation modeling (SEM) and causal inference methods, the latter having tools explicitly developed for decomposing a total causal effect into direct and indirect effects. The two main goals of the SEM model are to test the global hypothesis of whether our conceptual model fits the data (i.e., does our DAG conform to the observed associations) and obtain adjusted path estimates, which add statistical “weights” to the arrows in our DAG.

After making some data-driven modifications to the SEM model in the form of additional correlations, which can be thought of as undirected associations between nodes in the DAG, the SEM achieves good fit to the data per the usual fit indices. More on that below. The SEM parameter estimates suggest that ACE mainly acts upon BMI remission via the mediating variables, and not directly.

If all the variables in our DAG / SEM were continuous, and absent interactions between exposure and mediator, we would be able to use fairly straightforward product-of-coefficient methods to estimate the indirect effects. However, our main exposure (ACE) and outcome (BMI remission) are both binary, and several of the mediators are ordinal. Another complication arises from the **intermediate confounding** of alcohol on physical activity with respect to BMI remission, which means that alcohol is a confounder (common cause) of both the mediator and the outcome, but is also caused by ACE exposure itself and is therefore a mediator too. For technical reasons, this means that disentangling the direct and indirect effects among these variables is not easy, and in some cases not possible. For these two reasons, we need to use more advanced techniques, at least for the complex variables.

For certain mediators, we are able to use the product method, and those results are in the SEM section.

Analyses were performed with R version 4.3.1. The lavaan package is used to fit the SEM model, and the medflex package is used for joint mediation analysis. The R code used to import and clean the data is shown below, so you can see for yourself.

**Brief summary of results:** ACE has a significant total effect on BMI remission, adjusting for household income. The SEM suggests that the effect of ACE is mediated by Behaviour, Self Concept, and wave 1 (age 9) child BMI. The joint mediating pathway of alcohol and physical activity is not significant, per natural effects mediation analysis.

**Some comments about the data:**

- No missing data on our primary exposure, binary ACES (ACES_study_binary)
- N = 534 missing obs for child BMI category at wave 3 (w3_childBMI_CAT), and consequently the same amount of missing data on BMI remission at wave 3
- Family income was divided by 1000 so its variance isn’t vastly disproportionate to the other variables, which can cause problems with SEM model fit
- We start with N = 8136 total observations but end up with N = 2210 after subsetting to just those who are overweight or obese at baseline. When we fit the SEM model, we have approximately N = 1300 observations

*## importing main dataset*

GUI <- read.csv("C:\\Users\\dufaultb\\R PROGRAMS\\McGAVOCK\\SEERAT WARAICH\\GUI Sorted.csv")

*## dividing income by 1000 so its variance isn't vastly disproportionate to the other variables*

GUI <- GUI %>%

rename(w1_equivinc = w1_Equivinc) %>%

mutate(w1_equivinc = w1_equivinc / 1000,

w3_equivinc = w3_equivinc / 1000,

w3_childBMI_CAT = case_when(w3_childBMI_CAT == 999 ~ NA_real_,

TRUE ~ w3_childBMI_CAT) )

*## importing a comprehensive diet score at wave 2 (age 13) to replace "soda"*

diet.var <- read_excel(path = "C:\\Users\\dufaultb\\R PROGRAMS\\McGAVOCK\\Diet Quality Score Wave 2.xlsx") %>%

dplyr::select(id, Diet_Quality_Score_positivenegative) %>%

rename(ID = id,

diet_score = Diet_Quality_Score_positivenegative)

GUI <- left_join(GUI, diet.var, by = "ID")

*## keeping only those who are overweight or obese at baseline (wave 1, age 9)*

*## defining BMI remission as having BMI category = 1 (normal weight) at age 18*

GUI <- GUI %>%

filter(w1_BMI9z_CAT %**in**% c(2, 3)) %>%

mutate(BMI_remission = case_when(w3_childBMI_CAT == 1 ~ 1,

w3_childBMI_CAT %**in**% c(2, 3) ~ 0,

TRUE ~ NA_real_) )

*# converting '8' (refuse) and '9' (unknown) to missing values*

my.list <- c("w3_CQ_weekday_timeonline", "w3_CQ_weekend_timeonline", "w3_CQ_weekday_tv", "w3_CQ_weekend_tv",

"w3_CQ_weekday_videogames", "w3_CQ_weekend_videogames", "w3_CQ_multiscreen", "w3_CQ_smoked_cigarette",

"w3_CQ_alochol_FQ_b", "w3_CQ_exercise_past14days")

GUI <- GUI %>%

mutate(across(all_of(my.list), ~ case_when(. %**in**% c(8, 9) ~ NA_real_, TRUE ~ .)))

rm(diet.var, my.list)

Missing Data

This is a visual exploration of the missing data in the GUI subset, which includes only those who are overweight or obese at baseline (age 9). On the left-hand side, you can see the proportion missing for each variable. Most of them are moderate, with spikes for income, parental BMI, exercise (w3_CQ_exercise_past14days), and BMI remission. The worst by far, however, is smoking (w3_CQ_best_description_smoking), at about 60% missing, and therefore we lose *at least* that fraction of the sample for our multivariate analyses.

On the right-hand side we see a very informative plot showing the *patterns* of missing data, with blue tiles indicating that variable has been observed, and red tiles indicating it is missing. Each row is a pattern. The horizontal bar beside each row shows its frequency in the sample. The most frequent pattern in our data is shown at the bottom, and the least frequent is shown at the top. One main takeaways here is that the most common pattern is complete data, followed by those missing only cigarette use, followed again by those missing only income, parental BMI, exercise, cigarettes, alcohol, and BMI remission.

It’s interesting to note that the SDQ items and ACES have no missing data.

It may not be possible to include cigarettes in our SEM model, or any other multivariable analysis. If we use the version “have you ever smoked a cigarette?” (w3_CQ_smoked_cigarette) the percentage missing drops to about 20%, but this is still high and is a fairly uninformative measure of smoke exposure. We proceed without any measure of smoking, for now at least.

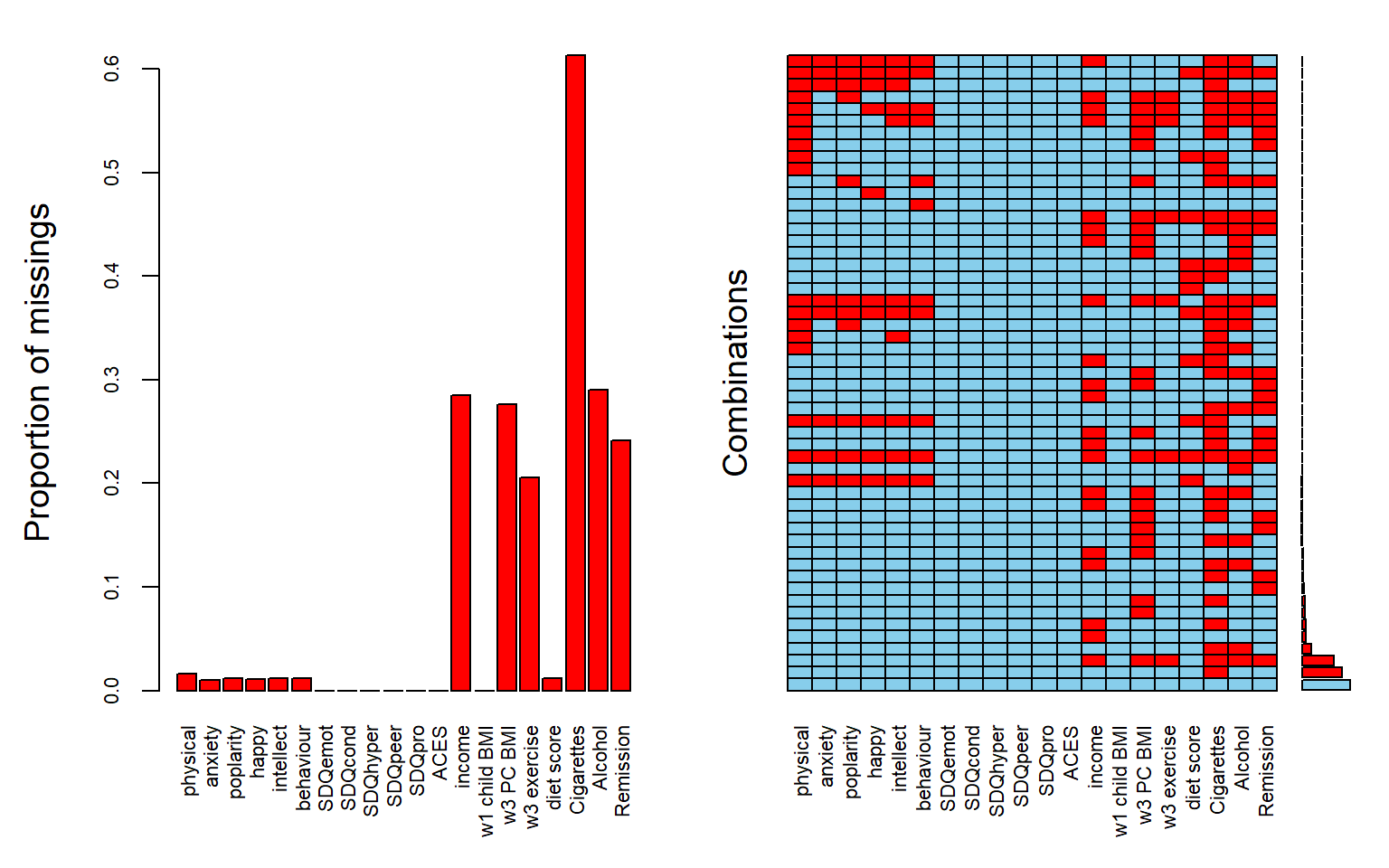

Summary Statistics

In this section we present summary statistics for the main variables involved in the SEM, plus a few others related to physical activity and demographics. The first two columns are the strata of ACE (yes vs no), and the rightmost column is the overall sample. This is for descriptive purposes.

|  | **No ACE (N=1917)** | **ACE (N=293)** | **Overall (N=2210)** |
| --- | --- | --- | --- |
| **Gender_Binary** |  |  |  |
| 1 | 969 (50.5%) | 121 (41.3%) | 1090 (49.3%) |
| 2 | 948 (49.5%) | 172 (58.7%) | 1120 (50.7%) |
| **w3_equivinc** |  |  |  |
| Mean (SD) | 16.2 (9.00) | 13.6 (6.19) | 15.8 (8.71) |
| Median [Min, Max] | 14.0 [5.00, 60.0] | 13.0 [5.00, 35.0] | 14.0 [5.00, 60.0] |
| Missing | 550 (28.7%) | 80 (27.3%) | 630 (28.5%) |
| **w3_PCG_BMI_MD** |  |  |  |
| Mean (SD) | 28.0 (5.24) | 28.1 (4.95) | 28.0 (5.20) |
| Median [Min, Max] | 27.0 [17.0, 47.0] | 27.0 [19.0, 43.0] | 27.0 [17.0, 47.0] |
| Missing | 524 (27.3%) | 87 (29.7%) | 611 (27.6%) |
| **w2_PCG_SDQ_emot** |  |  |  |
| Mean (SD) | 1.76 (1.94) | 2.56 (2.24) | 1.86 (2.00) |
| Median [Min, Max] | 1.00 [0, 10.0] | 2.00 [0, 10.0] | 1.00 [0, 10.0] |
| **w2_PCG_SDQ_cond** |  |  |  |
| Mean (SD) | 1.04 (1.33) | 1.51 (1.78) | 1.11 (1.40) |
| Median [Min, Max] | 1.00 [0, 10.0] | 1.00 [0, 9.00] | 1.00 [0, 10.0] |
| **w2_PCG_SDQ_hyper** |  |  |  |
| Mean (SD) | 2.47 (2.27) | 3.17 (2.62) | 2.56 (2.33) |
| Median [Min, Max] | 2.00 [0, 10.0] | 3.00 [0, 10.0] | 2.00 [0, 10.0] |
| **w2_PCG_SDQ_peer** |  |  |  |
| Mean (SD) | 1.12 (1.47) | 1.63 (1.83) | 1.19 (1.53) |
| Median [Min, Max] | 1.00 [0, 10.0] | 1.00 [0, 10.0] | 1.00 [0, 10.0] |
| **w2_PCG_SDQ_pro** |  |  |  |
| Mean (SD) | 8.94 (1.47) | 8.74 (1.61) | 8.91 (1.49) |
| Median [Min, Max] | 10.0 [0, 10.0] | 9.00 [3.00, 10.0] | 9.00 [0, 10.0] |
| **w2_PCG_SDQ_Tot** |  |  |  |
| Mean (SD) | 6.38 (4.96) | 8.87 (6.20) | 6.71 (5.21) |
| Median [Min, Max] | 5.00 [0, 35.0] | 8.00 [0, 29.0] | 6.00 [0, 35.0] |
| **w2_ph_behaviour** |  |  |  |
| Mean (SD) | 12.6 (2.10) | 12.1 (2.33) | 12.5 (2.13) |
| Median [Min, Max] | 13.0 [2.00, 14.0] | 13.0 [1.00, 14.0] | 13.0 [1.00, 14.0] |
| Missing | 20 (1.0%) | 6 (2.0%) | 26 (1.2%) |
| **w2_ph_intellectual** |  |  |  |
| Mean (SD) | 12.2 (3.04) | 11.4 (3.36) | 12.1 (3.10) |
| Median [Min, Max] | 13.0 [0, 16.0] | 12.0 [2.00, 16.0] | 13.0 [0, 16.0] |
| Missing | 21 (1.1%) | 6 (2.0%) | 27 (1.2%) |
| **w2_ph_physical** |  |  |  |
| Mean (SD) | 7.78 (2.41) | 7.25 (2.65) | 7.71 (2.44) |
| Median [Min, Max] | 8.00 [0, 11.0] | 8.00 [0, 11.0] | 8.00 [0, 11.0] |
| Missing | 30 (1.6%) | 6 (2.0%) | 36 (1.6%) |
| **w2_ph_free_anxiety** |  |  |  |
| Mean (SD) | 10.6 (3.10) | 9.75 (3.51) | 10.5 (3.17) |
| Median [Min, Max] | 11.0 [0, 14.0] | 11.0 [1.00, 14.0] | 11.0 [0, 14.0] |
| Missing | 20 (1.0%) | 3 (1.0%) | 23 (1.0%) |
| **w2_ph_popularity** |  |  |  |
| Mean (SD) | 9.68 (2.28) | 9.22 (2.53) | 9.62 (2.32) |
| Median [Min, Max] | 10.0 [0, 12.0] | 10.0 [0, 12.0] | 10.0 [0, 12.0] |
| Missing | 24 (1.3%) | 3 (1.0%) | 27 (1.2%) |
| **w2_ph_happiness** |  |  |  |
| Mean (SD) | 8.45 (1.74) | 8.01 (2.06) | 8.39 (1.80) |
| Median [Min, Max] | 9.00 [0, 10.0] | 9.00 [0, 10.0] | 9.00 [0, 10.0] |
| Missing | 21 (1.1%) | 4 (1.4%) | 25 (1.1%) |
| **w2_ph_totalscore** |  |  |  |
| Mean (SD) | 47.7 (8.46) | 45.1 (9.80) | 47.3 (8.69) |
| Median [Min, Max] | 50.0 [2.00, 60.0] | 47.0 [5.00, 59.0] | 49.0 [2.00, 60.0] |
| Missing | 28 (1.5%) | 7 (2.4%) | 35 (1.6%) |
| **w3_CQ_exercise_past14days** |  |  |  |
| 1 | 199 (10.4%) | 43 (14.7%) | 242 (11.0%) |
| 2 | 317 (16.5%) | 52 (17.7%) | 369 (16.7%) |
| 3 | 422 (22.0%) | 70 (23.9%) | 492 (22.3%) |
| 4 | 296 (15.4%) | 25 (8.5%) | 321 (14.5%) |
| 5 | 296 (15.4%) | 35 (11.9%) | 331 (15.0%) |
| Missing | 387 (20.2%) | 68 (23.2%) | 455 (20.6%) |
| **w3_CQ_cigaretteuse_week** |  |  |  |
| 0 | 73 (3.8%) | 10 (3.4%) | 83 (3.8%) |
| 1 | 72 (3.8%) | 11 (3.8%) | 83 (3.8%) |
| 2 | 23 (1.2%) | 6 (2.0%) | 29 (1.3%) |
| 3 | 33 (1.7%) | 13 (4.4%) | 46 (2.1%) |
| 4 | 19 (1.0%) | 7 (2.4%) | 26 (1.2%) |
| 5 | 17 (0.9%) | 11 (3.8%) | 28 (1.3%) |
| 6 | 10 (0.5%) | 2 (0.7%) | 12 (0.5%) |
| 7 | 24 (1.3%) | 17 (5.8%) | 41 (1.9%) |
| Missing | 1646 (85.9%) | 216 (73.7%) | 1862 (84.3%) |
| **w3_CQ_alochol_FQ_b** |  |  |  |
| 0 | 69 (3.6%) | 15 (5.1%) | 84 (3.8%) |
| 1 | 672 (35.1%) | 82 (28.0%) | 754 (34.1%) |
| 2 | 550 (28.7%) | 89 (30.4%) | 639 (28.9%) |
| 3 | 73 (3.8%) | 18 (6.1%) | 91 (4.1%) |
| Missing | 553 (28.8%) | 89 (30.4%) | 642 (29.0%) |
| **w1_child_BMI** |  |  |  |
| Mean (SD) | 21.1 (2.26) | 21.6 (2.36) | 21.2 (2.28) |
| Median [Min, Max] | 20.5 [18.2, 37.1] | 21.1 [18.3, 28.2] | 20.6 [18.2, 37.1] |
| **w1_BMI9z_CAT** |  |  |  |
| 2 | 1268 (66.1%) | 169 (57.7%) | 1437 (65.0%) |
| 3 | 649 (33.9%) | 124 (42.3%) | 773 (35.0%) |
| **w3_childBMI_CAT** |  |  |  |
| 1 | 687 (35.8%) | 84 (28.7%) | 771 (34.9%) |
| 2 | 535 (27.9%) | 91 (31.1%) | 626 (28.3%) |
| 3 | 239 (12.5%) | 40 (13.7%) | 279 (12.6%) |
| Missing | 456 (23.8%) | 78 (26.6%) | 534 (24.2%) |
| **w3_CQ_weekday_timeonline** |  |  |  |
| 1 | 28 (1.5%) | 9 (3.1%) | 37 (1.7%) |
| 2 | 160 (8.3%) | 16 (5.5%) | 176 (8.0%) |
| 3 | 444 (23.2%) | 48 (16.4%) | 492 (22.3%) |
| 4 | 347 (18.1%) | 45 (15.4%) | 392 (17.7%) |
| 5 | 347 (18.1%) | 64 (21.8%) | 411 (18.6%) |
| 6 | 181 (9.4%) | 40 (13.7%) | 221 (10.0%) |
| Missing | 410 (21.4%) | 71 (24.2%) | 481 (21.8%) |
| **w3_CQ_weekend_timeonline** |  |  |  |
| 1 | 20 (1.0%) | 7 (2.4%) | 27 (1.2%) |
| 2 | 82 (4.3%) | 7 (2.4%) | 89 (4.0%) |
| 3 | 235 (12.3%) | 22 (7.5%) | 257 (11.6%) |
| 4 | 397 (20.7%) | 44 (15.0%) | 441 (20.0%) |
| 5 | 589 (30.7%) | 93 (31.7%) | 682 (30.9%) |
| 6 | 184 (9.6%) | 49 (16.7%) | 233 (10.5%) |
| Missing | 410 (21.4%) | 71 (24.2%) | 481 (21.8%) |
| **w3_CQ_weekday_tv** |  |  |  |
| 1 | 214 (11.2%) | 30 (10.2%) | 244 (11.0%) |
| 2 | 522 (27.2%) | 73 (24.9%) | 595 (26.9%) |
| 3 | 453 (23.6%) | 70 (23.9%) | 523 (23.7%) |
| 4 | 192 (10.0%) | 28 (9.6%) | 220 (10.0%) |
| 5 | 81 (4.2%) | 12 (4.1%) | 93 (4.2%) |
| 6 | 45 (2.3%) | 9 (3.1%) | 54 (2.4%) |
| Missing | 410 (21.4%) | 71 (24.2%) | 481 (21.8%) |
| **w3_CQ_weekend_tv** |  |  |  |
| 1 | 107 (5.6%) | 19 (6.5%) | 126 (5.7%) |
| 2 | 237 (12.4%) | 42 (14.3%) | 279 (12.6%) |
| 3 | 485 (25.3%) | 66 (22.5%) | 551 (24.9%) |
| 4 | 437 (22.8%) | 52 (17.7%) | 489 (22.1%) |
| 5 | 189 (9.9%) | 29 (9.9%) | 218 (9.9%) |
| 6 | 52 (2.7%) | 14 (4.8%) | 66 (3.0%) |
| Missing | 410 (21.4%) | 71 (24.2%) | 481 (21.8%) |
| **w3_CQ_weekday_videogames** |  |  |  |
| 1 | 964 (50.3%) | 139 (47.4%) | 1103 (49.9%) |
| 2 | 242 (12.6%) | 34 (11.6%) | 276 (12.5%) |
| 3 | 136 (7.1%) | 19 (6.5%) | 155 (7.0%) |
| 4 | 74 (3.9%) | 14 (4.8%) | 88 (4.0%) |
| 5 | 63 (3.3%) | 11 (3.8%) | 74 (3.3%) |
| 6 | 28 (1.5%) | 4 (1.4%) | 32 (1.4%) |
| Missing | 410 (21.4%) | 72 (24.6%) | 482 (21.8%) |
| **w3_CQ_weekend_videogames** |  |  |  |
| 1 | 834 (43.5%) | 130 (44.4%) | 964 (43.6%) |
| 2 | 212 (11.1%) | 31 (10.6%) | 243 (11.0%) |
| 3 | 163 (8.5%) | 19 (6.5%) | 182 (8.2%) |
| 4 | 143 (7.5%) | 19 (6.5%) | 162 (7.3%) |
| 5 | 122 (6.4%) | 17 (5.8%) | 139 (6.3%) |
| 6 | 33 (1.7%) | 5 (1.7%) | 38 (1.7%) |
| Missing | 410 (21.4%) | 72 (24.6%) | 482 (21.8%) |
| **w3_CQ_multiscreen** |  |  |  |
| 1 | 513 (26.8%) | 77 (26.3%) | 590 (26.7%) |
| 2 | 317 (16.5%) | 37 (12.6%) | 354 (16.0%) |
| 3 | 307 (16.0%) | 47 (16.0%) | 354 (16.0%) |
| 4 | 225 (11.7%) | 30 (10.2%) | 255 (11.5%) |
| 5 | 144 (7.5%) | 30 (10.2%) | 174 (7.9%) |
| Missing | 411 (21.4%) | 72 (24.6%) | 483 (21.9%) |
| **BMI_remission** |  |  |  |
| 0 | 774 (40.4%) | 131 (44.7%) | 905 (41.0%) |
| 1 | 687 (35.8%) | 84 (28.7%) | 771 (34.9%) |
| Missing | 456 (23.8%) | 78 (26.6%) | 534 (24.2%) |

**BMI remission at age 18 by baseline weight status (overweight or obese)**

```
##
##
##    Cell Contents
## |-------------------------|
## |                       N |
## |           N / Row Total |
## |           N / Col Total |
## |-------------------------|
##
##
## Total Observations in Table:  1676
##
##
##              | BMI Remission
##       w1 BMI |         0 |         1 | Row Total |
## -------------|-----------|-----------|-----------|
##            2 |       489 |       618 |      1107 |
##              |     0.442 |     0.558 |     0.661 |
##              |     0.540 |     0.802 |           |
## -------------|-----------|-----------|-----------|
##            3 |       416 |       153 |       569 |
##              |     0.731 |     0.269 |     0.339 |
##              |     0.460 |     0.198 |           |
## -------------|-----------|-----------|-----------|
## Column Total |       905 |       771 |      1676 |
##              |     0.540 |     0.460 |           |
## -------------|-----------|-----------|-----------|
##
##
```

**Diet Score by ACE Status**

| **ACES_study_binary** | **N** | **min** | **mean** | **median** | **max** | **std.dev** |
| --- | --- | --- | --- | --- | --- | --- |
| 0 | 1893 | -13 | 1.923 | 2 | 14 | 3.729 |
| 1 | 291 | -10 | 1.546 | 1 | 10 | 3.916 |

```
##
##  Kruskal-Wallis rank sum test
##
## data:  GUI$diet_score by GUI$ACES_study_binary
## Kruskal-Wallis chi-squared = 2.76, df = 1, p-value = 0.09665
```

ACE and BMI Remission

In this section, we explore the simple but key question of whether childhood exposure to ACE has a significant **total effect** on the odds of BMI remission. This includes both the direct and indirect (mediated) effects. To provide an unbiased causal estimate of this effect, we adjust for the confounder of household income. Note that, per our DAG, this is the only measured common cause of ACE and BMI remission. I use wave 1 income instead of wave 3 income, but the results are similar for ACE either way.

Results of a logistic regression model with remission as the outcome are shown below. We see that being exposed to ACE reduces the odds of remission by approximately 25%, and is significant. Income is also significantly associated with remission; higher incomes increase the odds of remission by about 1% per thousand dollars.

Recall that the number of BMI remission events in our sample is 771, so this model has good power.

| **Logistic regression estimates:** | | | | |
| --- | --- | --- | --- | --- |
| **predictor** | **odds ratio** | **p.value** | **conf.low** | **conf.high** |
| (Intercept) | 0.7184 | 0.0011 | 0.5865 | 0.8729 |
| ACES_study_binary1 | 0.7328 | 0.0430 | 0.5407 | 0.9882 |
| w1_equivinc | 1.0102 | 0.0116 | 1.0026 | 1.0186 |

Structural Equation Model with Mediation

In this section we fit the full mediation model of interest, in which exposure to ACE at wave 1 is causative of BMI remission at wave 3 both directly and via indirect pathways involving behaviour, child BMI, and the latent factors captured by the PH and SDQ instruments. We also adjust for confounding by income where necessary.

As a reminder of what we are fitting, see the DAG sketch below. Note that you have hypothesized the existence of unmeasured environmental confounders.

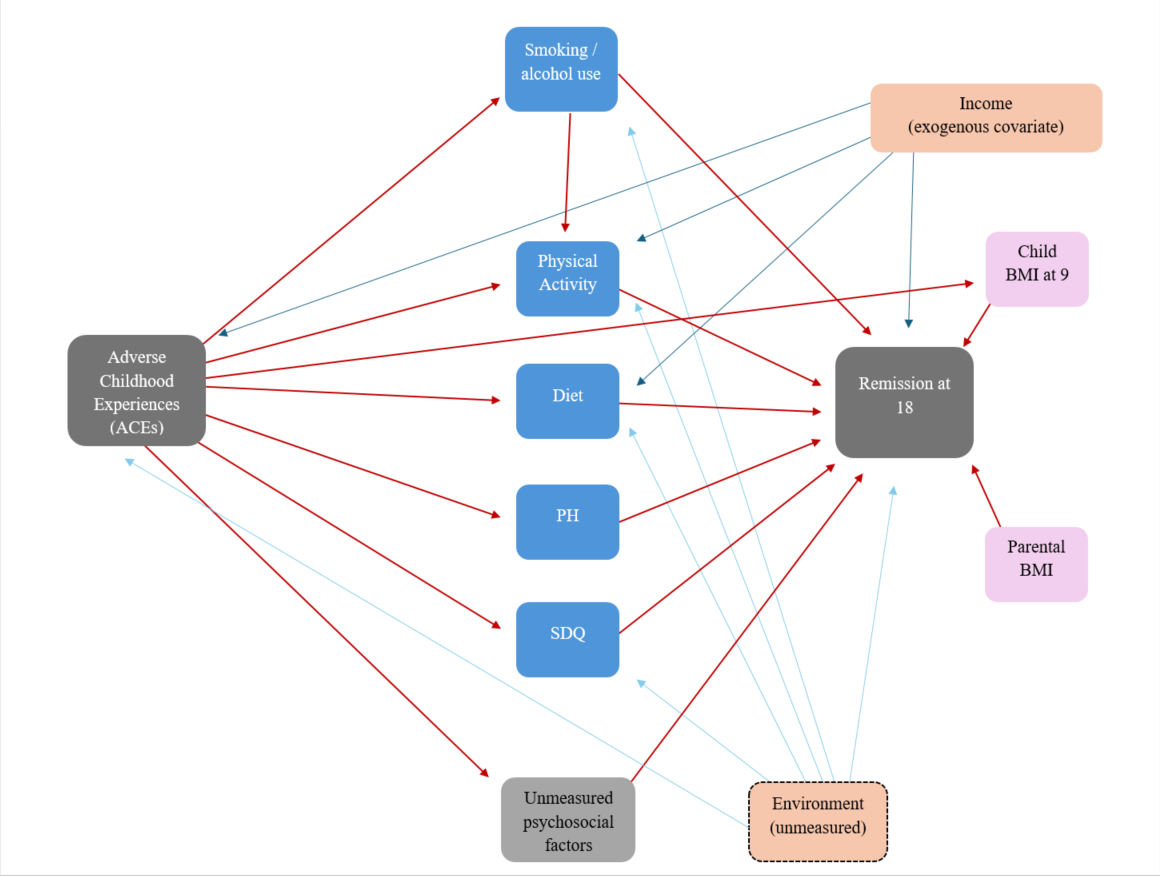

Hypothesized DAG Model

To obtain good fit statistics we added the correlations you see in the R code, including one between the Self Concept and Behaviour latent factors. We also allowed w2ph_behaviour to *cross-load* between them, meaning it is an indicator of both. Based on the name of the variable, it makes sense that it would also be an outcome of the Behaviour factor - but check the subscale content to verify. Again, these are data-driven changes to the model. There is no guarantee they make theoretical sense; you need to decide.

We used diagonally-weighted least squares (DWLS) because several of our outcomes, including ACES and BMI remission, are ordinal. We assume the observed ordinal scale is a proxy for an underlying continuum. This is an unavoidable assumption we must make to adopt the SEM framework, though in my opinion a sensible one. Though we actually are forced to go further and assume an underlying *normal* distribution, which may or may not be realistic. This is also known as an item response model. A quick sensitivity test in which we use robust ML but treat everything as continuous showed qualitatively similar results.

The “regressions” in the output below refer to any variable that has an arrow going into it. The “latent variables” are SDQ and PH. This is a theoretical advantage of the SEM approach: we can treat these variables as truly being latent, and account for measurement error and allow the subscores to have different weights / importance.

**Results:** we see that BMI remission is significantly predicted by exercise, Self Concept, Behaviour (albeit borderline), and wave 1 BMI. Not significant is ACE. However, this is only the direct path. ACE exposure is a significant predictor of all mediators except diet score. Informally, this suggests that the effect of ACE is mostly mediated by these variables.

The lavaan code is shown below, followed by the results.

*## Full SEM model*

*## Note that SMOKING has been omitted due to large amounts of missing data*

sem.model <- '## measurement models

Self_Concept =~ w2_ph_physical + w2_ph_free_anxiety + w2_ph_popularity +

w2_ph_happiness + w2_ph_intellectual + w2_ph_behaviour

Behaviour	=~ w2_PCG_SDQ_emot + w2_PCG_SDQ_peer + w2_PCG_SDQ_hyper + w2_PCG_SDQ_cond +

w2_PCG_SDQ_pro + w2_ph_behaviour

\## regressions

ACES_study_binary ~ w1_equivinc

w1_child_BMI ~ child1*ACES_study_binary

w3_CQ_alochol_FQ_b ~ ACES_study_binary

w3_CQ_exercise_past14days ~ ACES_study_binary + w1_equivinc + w3_CQ_alochol_FQ_b

diet_score	~ diet1*ACES_study_binary + w1_equivinc

Self_Concept ~ self1*ACES_study_binary

Behaviour	~ behv1*ACES_study_binary

BMI_remission ~ diet2*diet_score	+ w3_CQ_exercise_past14days +

self2*Self_Concept + behv2*Behaviour +

w3_CQ_alochol_FQ_b + w1_equivinc + ACES_study_binary +

child2*w1_child_BMI

\## + w3_PCG_BMI_MD

\## covariances

Self_Concept ~~ Behaviour

Self_Concept ~~ w3_CQ_exercise_past14days

Behaviour	~~ diet_score

Self_Concept ~~ diet_score

w2_ph_popularity ~~ w2_PCG_SDQ_peer

\# indirect effect via self-concept:

Indirect_Self := self1*self2

\# indirect effect via behaviour:

Indirect_Behv := behv1*behv2

\# indirect effect via diet score:

Indirect_Diet := diet1*diet2

\# indirect effect via wave 1 BMI:

Indirect_w1BMI := child1*child2 '

sem.fit <- sem(sem.model, data = GUI, std.lv = TRUE,

ordered = c("ACES_study_binary", "BMI_remission",

"w3_CQ_exercise_past14days", "w3_CQ_alochol_FQ_b"),

estimator = "DWLS", fixed.x = TRUE)

*## sem.fit <- sem(sem.model, data = GUI, std.lv = TRUE, estimator = "MLR", fixed.x = TRUE)*

mi.fit <- modificationindices(sem.fit, sort. = TRUE) %>% arrange(desc(mi))

And here is the model output, including final sample size, fit statistics, parameter estimates for path coefficients, latent loadings, and covariances

```
## lavaan 0.6-8 ended normally after 81 iterations
##
##   Estimator                                       DWLS
##   Optimization method                           NLMINB
##   Number of model parameters                        70
##
##                                                   Used       Total
##   Number of observations                          1376        2210
##
## Model Test User Model:
##
##   Test statistic                               603.620
##   Degrees of freedom                               118
##   P-value (Chi-square)                           0.000
##
## Model Test Baseline Model:
##
##   Test statistic                              8891.007
##   Degrees of freedom                               136
##   P-value                                        0.000
##
## User Model versus Baseline Model:
##
##   Comparative Fit Index (CFI)                    0.945
##   Tucker-Lewis Index (TLI)                       0.936
##
## Root Mean Square Error of Approximation:
##
##   RMSEA                                          0.055
##   90 Percent confidence interval - lower         0.050
##   90 Percent confidence interval - upper         0.059
##   P-value RMSEA <= 0.05                          0.035
##
## Standardized Root Mean Square Residual:
##
##   SRMR                                           0.052
##
## Parameter Estimates:
##
##   Standard errors                             Standard
##   Information                                 Expected
##   Information saturated (h1) model        Unstructured
##
## Latent Variables:
##                    Estimate  Std.Err  z-value  P(>|z|)   Std.lv  Std.all
##   Self_Concept =~
##     w2_ph_physical    1.941    0.050   38.847    0.000    1.963    0.791
##     w2_ph_fre_nxty    2.497    0.064   38.987    0.000    2.526    0.797
##     w2_ph_populrty    1.601    0.042   38.096    0.000    1.619    0.690
##     w2_ph_happinss    1.455    0.035   41.767    0.000    1.471    0.807
##     w2_ph_intllctl    2.408    0.059   41.032    0.000    2.436    0.793
##     w2_ph_behavior    0.772    0.036   21.371    0.000    0.781    0.388
##   Behaviour =~
##     w2_PCG_SDQ_emt    1.054    0.039   27.201    0.000    1.108    0.560
##     w2_PCG_SDQ_per    0.739    0.028   26.362    0.000    0.777    0.509
##     w2_PCG_SDQ_hyp    1.204    0.046   26.265    0.000    1.266    0.553
##     w2_PCG_SDQ_cnd    0.897    0.029   31.156    0.000    0.943    0.682
##     w2_PCG_SDQ_pro   -0.663    0.027  -24.741    0.000   -0.697   -0.468
##     w2_ph_behavior   -0.567    0.043  -13.166    0.000   -0.596   -0.296
##
## Regressions:
##                               Estimate  Std.Err  z-value  P(>|z|)   Std.lv
##   ACES_study_binary ~
##     w1_qvnc                     -0.015    0.002   -6.655    0.000   -0.015
##   w1_child_BMI ~
##     ACES_s_ (chl1)               0.474    0.065    7.292    0.000    0.474
##   w3_CQ_alochol_FQ_b ~
##     ACES_s_                      0.114    0.033    3.421    0.001    0.114
##   w3_CQ_exercise_past14days ~
##     ACES_s_                     -0.144    0.036   -3.947    0.000   -0.144
##     w1_qvnc                      0.003    0.002    1.506    0.132    0.003
##     w3_CQ__                      0.056    0.032    1.761    0.078    0.056
##   diet_score ~
##     ACES_s_ (dit1)              -0.201    0.138   -1.459    0.145   -0.201
##     w1_qvnc                      0.015    0.008    1.822    0.068    0.015
##   Self_Concept ~
##     ACES_s_ (slf1)              -0.148    0.021   -6.999    0.000   -0.146
##   Behaviour ~
##     ACES_s_ (bhv1)               0.317    0.032    9.887    0.000    0.302
##   BMI_remission ~
##     dit_scr (dit2)              -0.001    0.009   -0.061    0.952   -0.001
##     w3_CQ__                      0.108    0.039    2.789    0.005    0.108
##     Slf_Cnc (slf2)              -0.066    0.027   -2.468    0.014   -0.067
##     Behavir (bhv2)              -0.069    0.036   -1.893    0.058   -0.072
##     w3_CQ__                     -0.043    0.038   -1.114    0.265   -0.043
##     w1_qvnc                      0.004    0.002    1.591    0.112    0.004
##     ACES_s_                      0.031    0.066    0.472    0.637    0.031
##     w1__BMI (chl2)              -0.186    0.016  -11.743    0.000   -0.186
##   Std.all
##
##    -0.208
##
##     0.216
##
##     0.115
##
##    -0.145
##     0.044
##     0.056
##
##    -0.055
##     0.056
##
##    -0.150
##
##     0.308
##
##    -0.002
##     0.108
##    -0.066
##    -0.071
##    -0.042
##     0.052
##     0.031
##    -0.412
##
## Covariances:
##                       Estimate  Std.Err  z-value  P(>|z|)   Std.lv  Std.all
##  .Self_Concept ~~
##    .Behaviour           -0.294    0.015  -19.973    0.000   -0.294   -0.294
##    .w3_CQ_xrcs_p14       0.231    0.017   14.005    0.000    0.231    0.231
##  .Behaviour ~~
##    .diet_score          -0.579    0.093   -6.230    0.000   -0.579   -0.156
##  .Self_Concept ~~
##    .diet_score           0.436    0.058    7.502    0.000    0.436    0.117
##  .w2_ph_popularity ~~
##    .w2_PCG_SDQ_per      -0.636    0.091   -7.021    0.000   -0.636   -0.285
##
## Intercepts:
##                    Estimate  Std.Err  z-value  P(>|z|)   Std.lv  Std.all
##    .w2_ph_physical    7.499    0.134   56.074    0.000    7.499    3.022
##    .w2_ph_fre_nxty   10.221    0.189   54.201    0.000   10.221    3.226
##    .w2_ph_populrty    9.382    0.148   63.419    0.000    9.382    3.999
##    .w2_ph_happinss    8.300    0.107   77.267    0.000    8.300    4.551
##    .w2_ph_intllctl   11.760    0.189   62.322    0.000   11.760    3.830
##    .w2_ph_behavior   12.551    0.135   93.085    0.000   12.551    6.239
##    .w2_PCG_SDQ_emt    2.078    0.112   18.618    0.000    2.078    1.049
##    .w2_PCG_SDQ_per    1.299    0.084   15.543    0.000    1.299    0.851
##    .w2_PCG_SDQ_hyp    2.775    0.115   24.161    0.000    2.775    1.212
##    .w2_PCG_SDQ_cnd    1.212    0.079   15.304    0.000    1.212    0.877
##    .w2_PCG_SDQ_pro    9.026    0.084  108.094    0.000    9.026    6.061
##    .ACES_stdy_bnry    0.000                               0.000    0.000
##    .w1_child_BMI     21.321    0.106  201.414    0.000   21.321    9.518
##    .w3_CQ_lchl_FQ_    0.000                               0.000    0.000
##    .w3_CQ_xrcs_p14    0.000                               0.000    0.000
##    .diet_score        1.541    0.191    8.051    0.000    1.541    0.413
##    .BMI_remission     0.000                               0.000    0.000
##    .Self_Concept      0.000                               0.000    0.000
##    .Behaviour         0.000                               0.000    0.000
##
## Thresholds:
##                    Estimate  Std.Err  z-value  P(>|z|)   Std.lv  Std.all
##     ACES_stdy_bn|1    0.862    0.071   12.163    0.000    0.862    0.843
##     w3_CQ_lc_FQ_|1   -1.515    0.077  -19.719    0.000   -1.515   -1.505
##     w3_CQ_lc_FQ_|2    0.150    0.066    2.276    0.023    0.150    0.149
##     w3_CQ_lc_FQ_|3    1.685    0.077   21.945    0.000    1.685    1.674
##     w3_CQ_xrc_14|1   -1.035    0.060  -17.175    0.000   -1.035   -1.021
##     w3_CQ_xrc_14|2   -0.327    0.056   -5.876    0.000   -0.327   -0.322
##     w3_CQ_xrc_14|3    0.412    0.055    7.450    0.000    0.412    0.407
##     w3_CQ_xrc_14|4    0.971    0.058   16.640    0.000    0.971    0.958
##     BMI_remissn|t1   -3.748    0.344  -10.900    0.000   -3.748   -3.699
##
## Variances:
##                    Estimate  Std.Err  z-value  P(>|z|)   Std.lv  Std.all
##    .w2_ph_physical    2.303    0.349    6.608    0.000    2.303    0.374
##    .w2_ph_fre_nxty    3.659    0.577    6.342    0.000    3.659    0.365
##    .w2_ph_populrty    2.881    0.249   11.590    0.000    2.881    0.523
##    .w2_ph_happinss    1.161    0.163    7.122    0.000    1.161    0.349
##    .w2_ph_intllctl    3.498    0.480    7.280    0.000    3.498    0.371
##    .w2_ph_behavior    2.782    0.147   18.949    0.000    2.782    0.687
##    .w2_PCG_SDQ_emt    2.696    0.187   14.438    0.000    2.696    0.687
##    .w2_PCG_SDQ_per    1.725    0.086   20.089    0.000    1.725    0.741
##    .w2_PCG_SDQ_hyp    3.639    0.252   14.459    0.000    3.639    0.694
##    .w2_PCG_SDQ_cnd    1.021    0.079   12.959    0.000    1.021    0.535
##    .w2_PCG_SDQ_pro    1.731    0.086   20.041    0.000    1.731    0.781
##    .ACES_stdy_bnry    1.000                               1.000    0.957
##    .w1_child_BMI      4.783    0.173   27.718    0.000    4.783    0.953
##    .w3_CQ_lchl_FQ_    1.000                               1.000    0.987
##    .w3_CQ_xrcs_p14    1.000                               1.000    0.973
##    .diet_score       13.833    0.542   25.539    0.000   13.833    0.993
##    .BMI_remission     0.831                               0.831    0.809
##    .Self_Concept      1.000                               0.978    0.978
##    .Behaviour         1.000                               0.905    0.905
##
## Scales y*:
##                    Estimate  Std.Err  z-value  P(>|z|)   Std.lv  Std.all
##     ACES_stdy_bnry    1.000                               1.000    1.000
##     w3_CQ_lchl_FQ_    1.000                               1.000    1.000
##     w3_CQ_xrcs_p14    1.000                               1.000    1.000
##     BMI_remission     1.000                               1.000    1.000
##
## R-Square:
##                    Estimate
##     w2_ph_physical    0.626
##     w2_ph_fre_nxty    0.635
##     w2_ph_populrty    0.477
##     w2_ph_happinss    0.651
##     w2_ph_intllctl    0.629
##     w2_ph_behavior    0.313
##     w2_PCG_SDQ_emt    0.313
##     w2_PCG_SDQ_per    0.259
##     w2_PCG_SDQ_hyp    0.306
##     w2_PCG_SDQ_cnd    0.465
##     w2_PCG_SDQ_pro    0.219
##     ACES_stdy_bnry    0.043
##     w1_child_BMI      0.047
##     w3_CQ_lchl_FQ_    0.013
##     w3_CQ_xrcs_p14    0.027
##     diet_score        0.007
##     BMI_remission     0.191
##     Self_Concept      0.022
##     Behaviour         0.095
##
## Defined Parameters:
##                    Estimate  Std.Err  z-value  P(>|z|)   Std.lv  Std.all
##     Indirect_Self     0.010    0.004    2.290    0.022    0.010    0.010
##     Indirect_Behv    -0.022    0.012   -1.882    0.060   -0.022   -0.022
##     Indirect_Diet     0.000    0.002    0.060    0.952    0.000    0.000
##     Indirect_w1BMI   -0.088    0.015   -5.996    0.000   -0.088   -0.089
```

The fit statistics for the above model are good:

fitMeasures(sem.fit, c("cfi", "rmsea", "srmr"))

```
##   cfi rmsea  srmr
## 0.945 0.055 0.052
```

This table shows the estimated **covariances** (correlations). Please inspect them to make sure the direction of effect is sensible.

| **Variable 1** | **Variable 2** | **Correlation** | **pvalue** | **ci.lower** | **ci.upper** |
| --- | --- | --- | --- | --- | --- |
| Self_Concept | Behaviour | -0.2943 | 0 | -0.3232 | -0.2654 |
| Self_Concept | w3_CQ_exercise_past14days | 0.2312 | 0 | 0.1988 | 0.2635 |
| Behaviour | diet_score | -0.1556 | 0 | -0.2046 | -0.1066 |
| Self_Concept | diet_score | 0.1173 | 0 | 0.0865 | 0.1481 |
| w2_ph_popularity | w2_PCG_SDQ_peer | -0.2854 | 0 | -0.3673 | -0.2034 |

And finally, here are the standardized estimates of the **indirect effects** (mediated paths) between ACE, the stated mediator, and BMI remission. There is evidence that childhood BMI, Self Concept, and Behaviour significantly mediate ACE and BMI remission, but diet score does not. That said, it is not clear how to interpret the coefficients of the mediated paths, since they are on a linear scale. Recall that our SEM fits the data by assuming an underlying continuum for ACE and BMI remission.

| **Mediation Path** | **Formula** | **Estimate** | **pvalue** | **ci.lower** | **ci.upper** |
| --- | --- | --- | --- | --- | --- |
| Indirect_Self | self1*self2 | 0.0099 | 0.0214 | 0.0015 | 0.0184 |
| Indirect_Behv | behv1*behv2 | -0.0220 | 0.0604 | -0.0450 | 0.0010 |
| Indirect_Diet | diet1*diet2 | 0.0001 | 0.9518 | -0.0037 | 0.0039 |
| Indirect_w1BMI | child1*child2 | -0.0891 | 0.0000 | -0.1183 | -0.0599 |

Natural Effects Joint Mediation

As mentioned, the relationship between alcohol and physical activity adds a layer of complexity. This is because alcohol is a confounder but is also caused by exposure, which for technical reasons renders certain causal identities generally unidentifiable (i.e. they cannot be recovered from data), absent certain parametric assumptions. This scenario is known as “intermediate confounding”. An intuitive explanation of why this is problematic is that the natural direct effect of ACE with respect to physical activity is partially mediated by alcohol, but so is the indirect effect. The upshot is that this natural direct effect can’t easily be estimated.

One way of recovering some knowledge about the natural direct and indirect effects (NDE and NIE) in the presence of intermediate confounding is to ask a less granular question, and combine the paths going through at least one of the mediators, and possibly both of them, and call that the indirect path. This is known as **joint mediation**. We now undertake this analysis, focusing on just the relevant variables, not the full SEM. Since the other mediators are causally independent of alcohol and physical activity, no bias is introduced by leaving them out.

We use the medflex package to estimate the total, NDE, and NIE effects of ACE on BMI remission. Since we cannot use product-of-coefficient methods, we employ an imputation approach in which we impute counterfactuals based on observed relationships and then estimate the imputed data with a marginal structural model.

Results are shown below. We find that the indirect effect is small and highly non-significant. The direct effect and total effect are almost the same size therefore. So it appears that the relationship between ACE and BMI remission goes through paths other than [alcohol, activity]. The total effect estimated here is similar to that of our logistic regression above, but not significant. I wouldn’t put too much stock in this, since we have a smaller sample size here by virtue of including two extra variables (the mediators), and by using a different method of estimating standard errors.

These analyses are adjusted for wave 1 household income.

For more details on natural effects mediation models, please see [this article](https://cran.r-project.org/web/packages/medflex/vignettes/medflex.pdf)

```
## Natural effect model
## with robust standard errors based on the sandwich estimator
## ---
## Exposure: ACES_study_binary
## Mediator(s): w3_CQ_exercise_past14days, w3_CQ_alochol_FQ_b
## ---
## Parameter estimates:
##                      Estimate Std. Error z value Pr(>|z|)
## (Intercept)         -0.334571   0.114398  -2.925  0.00345 **
## ACES_study_binary01 -0.201024   0.160884  -1.249  0.21148
## ACES_study_binary11 -0.022943   0.024926  -0.920  0.35735
## w1_equivinc          0.009067   0.004647   1.951  0.05106 .
## ---
## Signif. codes:  0 '***' 0.001 '**' 0.01 '*' 0.05 '.' 0.1 ' ' 1
## Effect decomposition on the scale of the linear predictor
## ---
## conditional on: w1_equivinc
## with x* = 0, x = 1
## ---
##                         Estimate
## natural direct effect   -0.20102
## natural indirect effect -0.02294
## total effect            -0.22397
```

Here is a sort of forest plot figure showing the fitted natural effects mediation estimates on the odds ratio scale, along with their robust 95% confidence intervals. Note that the total effect OR is the product of the direct effect OR and the indirect effect OR.

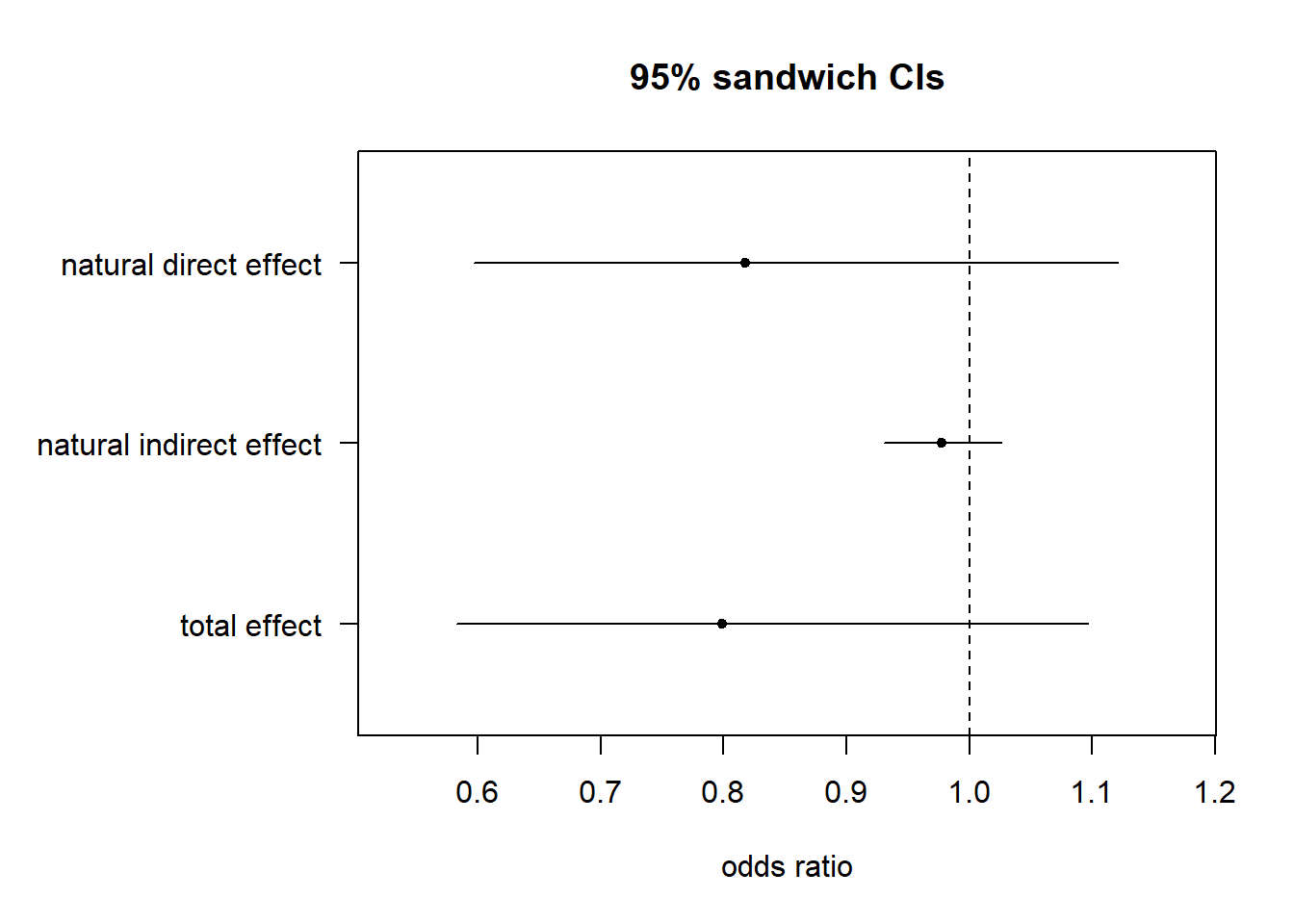

```
## Effect decomposition on exp(scale of the linear predictor)
## conditional on: w1_equivinc
## with x* = 0, x = 1
```

Table 1

This table provides the summary statistics and p-values for Table 1 of the manuscript. We have used the Fisher exact and Kruskal-Wallis tests to compare those with ACE versus those without. We included only those who met baseline inclusion criteria (overweight or obese at age 9) *and* had BMI (remission) recorded at age 18:

data = GUI %>%

filter(!is.na(BMI_remission) )

Note that I have omitted missing data from the descriptive statistics so that percentages would be more interpretable.

|  | **No ACE (N=1461)** | **ACE (N=215)** | **P-value** |
| --- | --- | --- | --- |
| **w1_BMI9z** |  |  |  |
| Mean (SD) | 1.81 (0.559) | 1.92 (0.621) | 0.0279 |
| Median [Min, Max] | 1.70 [1.02, 4.77] | 1.84 [1.04, 3.71] |  |
| **Gender_Binary** |  |  |  |
| 1 | 755 (51.7%) | 91 (42.3%) | 0.0106 |
| 2 | 706 (48.3%) | 124 (57.7%) |  |
| **w1_equivinc** |  |  |  |
| Mean (SD) | 21.4 (14.1) | 18.3 (11.9) | <0.001 |
| Median [Min, Max] | 18.9 [0.671, 216] | 15.4 [5.23, 120] |  |
| **w1_Measured_BMI_PCG** |  |  |  |
| Mean (SD) | 27.3 (4.95) | 26.9 (4.58) | 0.443 |
| Median [Min, Max] | 26.3 [16.9, 47.0] | 26.2 [18.1, 42.5] |  |
| **w1_hardexercise_past14days** |  |  |  |
| 1 | 31 (2.12%) | 7 (3.26%) | 0.602 |
| 2 | 89 (6.09%) | 15 (6.98%) |  |
| 3 | 297 (20.3%) | 37 (17.2%) |  |
| 4 | 316 (21.6%) | 50 (23.3%) |  |
| 5 | 728 (49.8%) | 106 (49.3%) |  |
| **diet_score** |  |  |  |
| Mean (SD) | 2.00 (3.70) | 1.71 (3.93) | 0.346 |
| Median [Min, Max] | 2.00 [-12.0, 14.0] | 2.00 [-9.00, 10.0] |  |
| **w1_PH_TotalScore** |  |  |  |
| Mean (SD) | 46.8 (8.31) | 45.0 (9.86) | 0.0548 |
| Median [Min, Max] | 48.0 [6.00, 60.0] | 48.0 [8.00, 59.0] |  |
| **w1_SDQ_total_PCG** |  |  |  |
| Mean (SD) | 7.48 (4.86) | 9.61 (5.95) | <0.001 |
| Median [Min, Max] | 7.00 [0, 28.0] | 8.00 [0, 31.0] |  |
| **w1_CESD_TOT_PCG** |  |  |  |
| Mean (SD) | 1.77 (2.85) | 3.41 (4.65) | <0.001 |
| Median [Min, Max] | 1.00 [0, 24.0] | 2.00 [0, 24.0] |  |

Table 2

This table provides the summary statistics and p-values for Table 2 of the manuscript. We have used the Fisher exact test to compare those with ACE versus those without. We included only those who met baseline inclusion criteria (overweight or obese at age 9) *and* had BMI (remission) recorded at age 18:

data = GUI %>%

filter(!is.na(BMI_remission) )

Note that these are **wave 3** variables being compared between ACE groups. Note also that I have omitted missing data from the descriptive statistics so that percentages would be more interpretable.

The raw data values for w3_CQ_age_smoked are “1” and then “13” to “18”, plus codes for missing response. I have grouped 13-18 and will let you decide whether “1” is 12 and under, as I think was assumed in your original table. Some variables had a few “8” responses that were treated as missing.

|  | **No ACE (N=1461)** | **ACE (N=215)** | **P-value** |
| --- | --- | --- | --- |
| **w3_CQ_exercise_past14days** |  |  |  |
| 1 | 181 (12.4%) | 39 (18.1%) | 0.006 |
| 2 | 303 (20.7%) | 49 (22.8%) |  |
| 3 | 407 (27.9%) | 69 (32.1%) |  |
| 4 | 283 (19.4%) | 24 (11.2%) |  |
| 5 | 287 (19.6%) | 34 (15.8%) |  |
| **w3_CQ_smoked_cigarette** |  |  |  |
| 1 | 688 (47.7%) | 128 (59.8%) | 0.00123 |
| 2 | 753 (52.3%) | 86 (40.2%) |  |
| **w3_CQ_age_smoked** |  |  |  |
| 1 | 48 (7.05%) | 18 (14.1%) | 0.0127 |
| 13 - 18 | 633 (93.0%) | 110 (85.9%) |  |
| **w3_CQ_cigaretteuse_week** |  |  |  |
| 0 - 4 | 210 (82.0%) | 45 (60.0%) | <0.001 |
| 5 + | 46 (18.0%) | 30 (40.0%) |  |
| **w3_CQ_ecigar_vaping** |  |  |  |
| 1 | 463 (32.1%) | 91 (42.5%) | 0.00313 |
| 2 | 979 (67.9%) | 123 (57.5%) |  |
| **w3_CQ_alochol_consum** |  |  |  |
| 1 | 1303 (90.4%) | 195 (91.1%) | 0.804 |
| 2 | 138 (9.58%) | 19 (8.88%) |  |
| **w3_Since13_Divorce** |  |  |  |
| 0 | 1370 (93.8%) | 156 (73.6%) | <0.001 |
| 1 | 91 (6.23%) | 56 (26.4%) |  |
| **w3_Since13_Violence** |  |  |  |
| 0 | 1344 (92.0%) | 173 (81.6%) | <0.001 |
| 1 | 117 (8.01%) | 39 (18.4%) |  |
| **w3_Since13_LostBestfriend** |  |  |  |
| 0 | 1361 (93.2%) | 189 (88.7%) | 0.0253 |
| 1 | 100 (6.84%) | 24 (11.3%) |  |

Table 3

This table provides the summary statistics and p-values for Table 3 of the manuscript. We have used the Kruskall-Wallis non-parametric test to compare those with ACE versus those without. As above, we included only those who met baseline inclusion criteria (overweight or obese at age 9) *and* had BMI (remission) recorded at age 18:

data = GUI %>%

filter(!is.na(BMI_remission) )

Note that these are **wave 2** variables being compared between ACE groups. Note also the missing data, which is mainly tiny but for w2_Depres_tot_SCG.

|  | **No ACE (N=1461)** | **ACE (N=215)** | **P-value** |
| --- | --- | --- | --- |
| **w2_ph_behaviour** |  |  |  |
| Mean (SD) | 12.7 (1.97) | 12.2 (2.28) | <0.001 |
| Median [Min, Max] | 13.0 [2.00, 14.0] | 13.0 [1.00, 14.0] |  |
| Missing | 13 (0.9%) | 2 (0.9%) |  |
| **w2_ph_intellectual** |  |  |  |
| Mean (SD) | 12.3 (2.96) | 11.3 (3.41) | <0.001 |
| Median [Min, Max] | 13.0 [0, 16.0] | 12.0 [2.00, 16.0] |  |
| Missing | 15 (1.0%) | 2 (0.9%) |  |
| **w2_ph_physical** |  |  |  |
| Mean (SD) | 7.79 (2.40) | 7.20 (2.75) | 0.00777 |
| Median [Min, Max] | 8.00 [0, 11.0] | 8.00 [0, 11.0] |  |
| Missing | 21 (1.4%) | 2 (0.9%) |  |
| **w2_ph_free_anxiety** |  |  |  |
| Mean (SD) | 10.5 (3.10) | 9.69 (3.55) | 0.00213 |
| Median [Min, Max] | 11.0 [0, 14.0] | 11.0 [1.00, 14.0] |  |
| Missing | 14 (1.0%) | 1 (0.5%) |  |
| **w2_ph_popularity** |  |  |  |
| Mean (SD) | 9.63 (2.29) | 9.24 (2.61) | 0.059 |
| Median [Min, Max] | 10.0 [0, 12.0] | 10.0 [0, 12.0] |  |
| Missing | 16 (1.1%) | 1 (0.5%) |  |
| **w2_ph_happiness** |  |  |  |
| Mean (SD) | 8.45 (1.73) | 7.95 (2.15) | 0.00483 |
| Median [Min, Max] | 9.00 [0, 10.0] | 9.00 [0, 10.0] |  |
| Missing | 15 (1.0%) | 1 (0.5%) |  |
| **w2_ph_totalscore** |  |  |  |
| Mean (SD) | 47.7 (8.33) | 45.0 (10.1) | <0.001 |
| Median [Min, Max] | 50.0 [2.00, 60.0] | 47.0 [5.00, 59.0] |  |
| Missing | 20 (1.4%) | 3 (1.4%) |  |
| **w2_PCG_SDQ_emot** |  |  |  |
| Mean (SD) | 1.75 (1.94) | 2.52 (2.14) | <0.001 |
| Median [Min, Max] | 1.00 [0, 10.0] | 2.00 [0, 10.0] |  |
| **w2_PCG_SDQ_cond** |  |  |  |
| Mean (SD) | 1.00 (1.30) | 1.45 (1.77) | <0.001 |
| Median [Min, Max] | 1.00 [0, 10.0] | 1.00 [0, 9.00] |  |
| **w2_PCG_SDQ_hyper** |  |  |  |
| Mean (SD) | 2.40 (2.23) | 3.11 (2.63) | <0.001 |
| Median [Min, Max] | 2.00 [0, 10.0] | 3.00 [0, 10.0] |  |
| **w2_PCG_SDQ_peer** |  |  |  |
| Mean (SD) | 1.12 (1.48) | 1.68 (1.86) | <0.001 |
| Median [Min, Max] | 1.00 [0, 10.0] | 1.00 [0, 8.00] |  |
| **w2_PCG_SDQ_pro** |  |  |  |
| Mean (SD) | 8.96 (1.45) | 8.68 (1.62) | 0.0147 |
| Median [Min, Max] | 10.0 [0, 10.0] | 9.00 [3.00, 10.0] |  |
| **w2_PCG_SDQ_Tot** |  |  |  |
| Mean (SD) | 6.28 (4.89) | 8.76 (6.14) | <0.001 |
| Median [Min, Max] | 5.00 [0, 35.0] | 8.00 [0, 27.0] |  |
| **w2_Depres_tot_PCG** |  |  |  |
| Mean (SD) | 2.13 (3.01) | 4.31 (5.38) | <0.001 |
| Median [Min, Max] | 1.00 [0, 23.0] | 3.00 [0, 24.0] |  |
| Missing | 10 (0.7%) | 2 (0.9%) |  |
| **w2_Depres_tot_SCG** |  |  |  |
| Mean (SD) | 1.49 (2.52) | 2.22 (3.45) | 0.392 |
| Median [Min, Max] | 1.00 [0, 22.0] | 0 [0, 16.0] |  |
| Missing | 268 (18.3%) | 141 (65.6%) |  |

Table 4

This table provides the summary statistics and p-values for Table 4 of the manuscript. We have used the Kruskall-Wallis non-parametric test to compare those with ACE versus those without. As above, we included only those who met baseline inclusion criteria (overweight or obese at age 9) *and* had BMI (remission) recorded at age 18:

data = GUI %>%

filter(!is.na(BMI_remission) )

All of these variables are from wave 3. Note that my variable names differ from what seems to be in your data dictionary, but the descriptions match (please double check). The income variable w3_equivinc has been divided by 1,000 to improve convergence in the SEM model, but you can change the scale as you wish; the p-value will be the same regardless.

|  | **No ACE (N=1461)** | **ACE (N=215)** | **P-value** |
| --- | --- | --- | --- |
| **w3_CQ_TIPI_extravert** |  |  |  |
| Mean (SD) | 4.81 (1.36) | 4.69 (1.35) | 0.305 |
| Median [Min, Max] | 5.00 [1.00, 7.00] | 5.00 [1.00, 7.00] |  |
| Missing | 2 (0.1%) | 0 (0%) |  |
| **w3_CQ_TIPI_agreeable** |  |  |  |
| Mean (SD) | 4.70 (1.01) | 4.70 (1.14) | 0.777 |
| Median [Min, Max] | 4.50 [1.00, 7.00] | 4.50 [1.50, 7.00] |  |
| Missing | 3 (0.2%) | 0 (0%) |  |
| **w3_CQ_TIPI_conscientious** |  |  |  |
| Mean (SD) | 5.20 (1.13) | 5.12 (1.25) | 0.391 |
| Median [Min, Max] | 5.50 [1.50, 7.00] | 5.00 [1.00, 7.00] |  |
| Missing | 2 (0.1%) | 0 (0%) |  |
| **w3_CQ_TIPI_openess** |  |  |  |
| Mean (SD) | 5.49 (1.02) | 5.63 (0.961) | 0.08 |
| Median [Min, Max] | 5.50 [1.00, 7.00] | 6.00 [2.50, 7.00] |  |
| Missing | 2 (0.1%) | 0 (0%) |  |
| **w3_CQ_TIPI_emostabi** |  |  |  |
| Mean (SD) | 4.77 (1.34) | 4.46 (1.48) | 0.00305 |
| Median [Min, Max] | 5.00 [1.00, 7.00] | 4.50 [1.00, 7.00] |  |
| Missing | 2 (0.1%) | 0 (0%) |  |
| **w3_CQ_sg2_control** |  |  |  |
| Mean (SD) | 3.40 (0.687) | 3.21 (0.728) | <0.001 |
| Median [Min, Max] | 3.50 [1.00, 5.00] | 3.30 [1.00, 5.00] |  |
| Missing | 24 (1.6%) | 7 (3.3%) |  |
| **w3_CQ_CSI_probsolving** |  |  |  |
| Mean (SD) | 16.6 (5.10) | 15.9 (4.86) | 0.0446 |
| Median [Min, Max] | 16.0 [5.00, 30.0] | 16.0 [5.00, 29.0] |  |
| Missing | 26 (1.8%) | 4 (1.9%) |  |
| **w3_CQ_CSI_avoidance** |  |  |  |
| Mean (SD) | 13.5 (5.46) | 15.4 (6.05) | <0.001 |
| Median [Min, Max] | 13.0 [6.00, 36.0] | 15.0 [6.00, 36.0] |  |
| Missing | 26 (1.8%) | 4 (1.9%) |  |
| **w3_CQ_selfesteem_total** |  |  |  |
| Mean (SD) | 12.0 (3.50) | 10.9 (3.98) | <0.001 |
| Median [Min, Max] | 12.0 [0, 18.0] | 11.0 [1.00, 18.0] |  |
| Missing | 23 (1.6%) | 4 (1.9%) |  |
| **w3_CQ_ILCtot** |  |  |  |
| Mean (SD) | 23.8 (3.08) | 24.0 (3.47) | 0.252 |
| Median [Min, Max] | 24.0 [7.00, 30.0] | 24.0 [5.00, 30.0] |  |
| Missing | 366 (25.1%) | 45 (20.9%) |  |
| **w3_CES_tot_PCG** |  |  |  |
| Mean (SD) | 2.43 (3.35) | 4.23 (5.07) | <0.001 |
| Median [Min, Max] | 1.00 [0, 24.0] | 2.00 [0, 24.0] |  |
| Missing | 32 (2.2%) | 3 (1.4%) |  |
| **w3_PCG_TIP_stress** |  |  |  |
| Mean (SD) | 10.1 (3.71) | 11.9 (4.34) | <0.001 |
| Median [Min, Max] | 9.00 [6.00, 30.0] | 11.0 [6.00, 30.0] |  |
| Missing | 35 (2.4%) | 4 (1.9%) |  |
| **w3_equivinc** |  |  |  |
| Mean (SD) | 16.3 (9.02) | 13.7 (6.19) | <0.001 |
| Median [Min, Max] | 14.0 [5.00, 60.0] | 13.0 [5.00, 35.0] |  |
| Missing | 149 (10.2%) | 11 (5.1%) |  |

Parametric G-computation

Here, we simulate the causal contrast E[Y(1,M1(1),M2(0,M1(0))−Y(1,M1(0),M2(0,M1(0))]E[Y(1,M1(1),M2(0,M1(0))−Y(1,M1(0),M2(0,M1(0))], which is the path through M1M1 only. This is part of the natural indirect effect. Each component is estimated separately, and then we take the difference as the point estimate.

To generate confidence intervals, this process needs to be done repeatedly via bootstrap.

*## Create a copy of the original data for counterfactual scenarios*

data_cf <-

GUI %>%

filter(!is.na(ACES_study_binary),

!is.na(w3_CQ_alochol_FQ_b),

!is.na(w3_CQ_exercise_past14days),

!is.na(BMI_remission) ) %>%

dplyr::select(ID, ACES_study_binary, w3_CQ_alochol_FQ_b,

w3_CQ_exercise_past14days, BMI_remission) %>%

mutate(ACES_study_binary = as.factor(ACES_study_binary))

*## ------------------------------------- ##*

*## Step 1: Fit models for M1, M2, and Y*

*## ------------------------------------- ##*

model_M1 <- glm(w3_CQ_alochol_FQ_b ~ ACES_study_binary,

data = data_cf, family = "gaussian")

model_M2 <- glm(w3_CQ_exercise_past14days ~ ACES_study_binary + w3_CQ_alochol_FQ_b,

data = data_cf, family = "gaussian")

model_Y <- glm(BMI_remission ~ ACES_study_binary + w3_CQ_alochol_FQ_b + w3_CQ_exercise_past14days,

data = data_cf, family = "binomial")

*## --------------------------------------- ##*

*# Step 2: Simulate counterfactual outcomes*

*## --------------------------------------- ##*

set.seed(123) *# For reproducibility*

*# Simulate M1 under X=1 and X=0*

M1_1 <- rnorm(nrow(data_cf),

mean = predict(model_M1, newdata = data.frame(ACES_study_binary = "1"),

type = "response"),

sd	= sqrt(summary(model_M1)$dispersion))

M1_0 <- rnorm(nrow(data_cf),

mean = predict(model_M1, newdata = data.frame(ACES_study_binary = "0"),

type = "response"),

sd	= sqrt(summary(model_M1)$dispersion))

*# Simulate M2 under X=0 and M1_0*

M2_0_M1_0 <- rnorm(nrow(data_cf),

mean = predict(model_M2,

newdata = data.frame(

ACES_study_binary = "0",

w3_CQ_alochol_FQ_b = M1_0),

type = "response"),

sd = sqrt(summary(model_M2)$dispersion))

*# Simulate Y for the contrast*

*# Y(1, M1(1), M2(0, M1(0)))*

prob_Y_1 <- predict(model_Y,

newdata = data.frame(ACES_study_binary = "1",

w3_CQ_alochol_FQ_b = M1_1,

w3_CQ_exercise_past14days = M2_0_M1_0),

type = "response")

Y_1_M1_1_M2_0_M1_0 <- rbinom(nrow(data_cf), size = 1, prob = prob_Y_1)

*# Y(1, M1(0), M2(0, M1(0)))*

prob_Y_2 <- predict(model_Y,

newdata = data.frame(ACES_study_binary = "1",

w3_CQ_alochol_FQ_b = M1_0,

w3_CQ_exercise_past14days = M2_0_M1_0),

type = "response")

Y_1_M1_0_M2_0_M1_0 <- rbinom(nrow(data_cf), size = 1, prob = prob_Y_2)

*## ------------------------------------- ##*

*## Step 3: Calculate the NIE via M1*

*## ------------------------------------- ##*

NIE_via_M1 <- mean(Y_1_M1_1_M2_0_M1_0 - Y_1_M1_0_M2_0_M1_0)

*# Output the result*

cat("Estimated Natural Indirect Effect (via M1) for binary Y:", NIE_via_M1, "\n")
